# The challenges of caring for people dying from COVID-19: a multinational, observational study of palliative and hospice services (CovPall)

**DOI:** 10.1101/2020.10.30.20221465

**Authors:** AO Oluyase, M Hocaoglu, R Cripps, M Maddocks, C Walshe, LK Fraser, N Preston, L Dunleavy, A Bradshaw, FEM Murtagh, S Bajwah, KE Sleeman, IJ Higginson, On behalf of the CovPall study team

**Author notes:** Corresponding author: Professor Irene J Higginson Address: Cicely Saunders Institute of Palliative Care, Policy and Rehabilitation, Florence Nightingale Faculty of Nursing, Midwifery & Palliative Care, Bessemer Road, London, SE5 9PJ, UK, |.

## Abstract

**Background:** Systematic data on the care of people dying with COVID-19 are scarce. We studied the response of and challenges for palliative care services during the COVID-19 pandemic.

**Methods:** We surveyed palliative care and hospice services, contacted via relevant organisations. Multivariable logistic regression identified associations with key challenges. Content analysis explored free text.

**Findings:** 458 services responded; 277 UK, 85 rest of Europe, 95 rest of the world (1 country unreported); 81% cared for patients with suspected or confirmed COVID-19, 77% had staff with suspected or confirmed COVID-19; 48% reported shortages of Personal Protective Equipment (PPE), 40% staff shortages, 24% medicines shortages, 14% shortages of other equipment. Services provided direct care and education in symptom management and communication; 91% changed how they worked. Care often shifted to increased community and hospital care, with fewer admissions to inpatient palliative care units. Factors associated with increased odds of PPE shortages were: charity rather than public management (OR 3·07, 95% CI 1·81-5·20), inpatient palliative care unit rather than other setting (OR 2·34, 95% CI 1·46-3·75). Being outside the UK was associated with lower odds of staff shortages (OR 0·44, 95% CI 0·26-0·76). Staff described increased workload, concerns for their colleagues who were ill, whilst expending time struggling to get essential equipment and medicines, perceiving they were not a front-line service.

**Interpretation:** Across all settings palliative care services were often overwhelmed, yet felt ignored in the COVID-19 response. Palliative care needs better integration with health care systems when planning and responding to future epidemics/pandemics.

**Funding:** MRC grant number MR/V012908/1, Cicely Saunders International and NIHR ARC South London.

**Research in context:** *Evidence before this study:* Systematic data on the response of palliative care services during COVID-19 are lacking. A search of PubMed on 27 August 2020 (start date: 01 December 2019) using keywords (palliative care OR end of life care OR hospice) and (COVID-19 OR coronavirus) and (multinational OR international) identified no studies that reported multinational or international data; there were 79 articles, mostly opinion pieces, single centre case studies or reports. A search for systematic reviews about palliative care and hospice services during pandemics of PubMed, with the same time periods and the keywords (palliative care OR end of life care OR hospice) and (COVID-19 OR coronavirus OR SARS-CoV-2) and (systematic review OR meta-analysis), identified one systematic review by Etkind et al, which underpinned this research and shares two senior authors (Higginson, Sleeman). Of 3094 articles identified, 10 studies, all observational, considered the palliative care response in pandemics. Studies were from single units or countries: West Africa, Taiwan, Hong Kong, Singapore, the U.S. (a simulation), and Italy (the only one considering COVID-19). The review concluded hospice and palliative care services are essential in the response to COVID-19 but systematic data are urgently needed to inform how to improve care for those who are likely to die, and/or have severe symptoms.

*Added value of this study:* We found a high response by palliative care services during the COVID-19 pandemic. Services cared for a surge in patients dying from and with severe symptoms due to COVID-19 in three main categories: patients with underlying conditions and/or multimorbid disease not previously known to palliative care (70% of services), patients already known to palliative care services (47% of services), and patients, previously healthy, now dying from COVID-19 (37% of services). More than three quarters of services reported having staff with suspected or confirmed COVID-19. We found high levels of shortages of Personal Protective Equipment (PPE), staff, medicines and other equipment, with different effects according to service management, care settings and world regions. Mitigating these challenges was extremely time consuming, limiting the palliative care response.

*Implications of all the available evidence:* Despite actively supporting dying patients, those with severe symptoms, their families/carers, and supporting other clinicians, palliative care professionals felt ignored by national health systems during the COVID-19 pandemic. Palliative care services need equipment, medicines and adequate staff to contribute fully to the pandemic response. Their crucial role must be better recognised and integrated, including into infection disease management, with improved workforce planning and management, so that patients and families can be better supported.

## Introduction

COVID-19 evolved from a mystery illness to a pandemic in 93 days, overwhelming health services in many countries.^1^ The World Health Organisation rapidly issued guidance on maintaining essential health services during the pandemic, highlighting prevention, maternal care, emergency care and chronic diseases, but without mention of palliative care.^2^ Yet COVID-19 has an overall case fatality ratio estimated between 1 and 4%;^3^ people with multimorbidity are at high risk of serious illness and death.^4^ By October 2020 there were over a million confirmed COVID-19 deaths worldwide.^5^

Palliative and hospice care (referred to hereafter as palliative care) is multidisciplinary, holistic and person-centred treatment, care and support for people with life-limiting illness, and those important to them, such as family and friends. Palliative care delivers expert pain and symptom control, care of dying patients, psychosocial support for patients, carers and health care professionals, supports triage and complex decision making.^6^ It is recommended as part of infectious disease care for older people,^7^ those with HIV/AIDS^8^ and drug resistant tuberculosis.^9^ Palliative care services, including hospices, support patients wherever they want to be cared for^10^ (see box 1). A Lancet editorial recommended that providing high-quality palliative care, with access to essential medicines, is vital during pandemics.^1,2^

### Box 1 Palliative care services provided in different settings

Palliative care and hospice services have multiprofessional teams of dedicated staff trained in palliative care, comprising doctors, nurses, and often social workers and therapists. They provide expertise in pain and symptom management, holistic and psychosocial care, decision making, advance care planning, end of life care and often bereavement support. They include support in the following settings (one service may provide support in one or more setting):

1. Inpatient palliative care unit – provides specialist inpatient palliative care. It can be a ward within, or adjacent to, a hospital, or a free standing building. In some countries, it is called an inpatient hospice.
2. Hospital palliative care team – provides specialist palliative care advice and support to other clinical staff, patients and their families and carers in the hospital environment. They offer formal and informal education and liaise with other services in and out of the hospital.
3. Home palliative care team – provides specialist palliative care to patients who need it at home, or in care homes or residential homes, and support their families and carers. They also provide specialist advice to general practitioners, family doctors and nurses caring for the patients.
4. Home nursing – provides intensive home nursing care for the patient at home, sometimes referred to as hospice or hospital at home, often supporting patients whose care needs are such that without this they would be admitted to an inpatient palliative care unit or hospital.

Palliative care services are often managed separately from other medical services and may be exceptionally vulnerable to disruption in pandemics as they often rely on charity funding. During the COVID-19 pandemic there were media reports of acute shortages of Personal Protective Equipment (PPE) and medicines that limited care.^11^ There is little systematic data about palliative care in these situations.^12,13^ This study aimed to understand the response of and challenges faced by palliative care services during the COVID-19 pandemic, and to identify factors associated with challenges experienced, in particular shortages of equipment, medicines and staff. We tested two a priori null hypotheses:

- there are no differences in shortages between services with different management type (e.g. charity and public)
- there are no differences in shortages between settings; e.g. between hospital based and non-hospital based; or community and non-community settings.

## Methods

### Study design and participants

CovPall is a multicentre multinational observational study of palliative care during the COVID-19 pandemic. This paper reports an on-line survey of palliative care services, the first main component of CovPall. The survey received ethical (Institutional Review Board) approval from King’s College London Research Ethics committee (LRS-19/20-18541); study sponsor: King’s College London, co-sponsor: King’s College Hospital NHS Foundation Trust, registered ISRCTN 16561225. It is reported according to CHERRIES,^14^ STROBE^15^ and MORECARE^16^ statements.

Inclusion criteria: Any palliative care service (box 1)^10^; caring for adults, children or both; managed by charity, public, private or other sector. Exclusion criteria: not a specialist palliative care/hospice service, i.e. having no members of staff with specific expertise/training in palliative care.

### Procedures and questionnaire

Services were identified and contacted through national and multinational palliative care and hospice organisations (supplementary file, box S1) and provided with a link to complete an on-line survey. A participant information sheet accompanied the invitation. Completion indicated consent. The questionnaire was developed and piloted by the CovPall team building on an earlier survey of Italian hospices,^12^ adding questions on the impact of and response to COVID-19. It was intended to be brief, taking about 30 minutes to complete. Free-text explanatory comments were invited. Data were anonymised before analysis.

### Analysis

After removing duplicate and ineligible entries all available data were analysed. We report completion rate and summary statistics. Missing data were not imputed. We used contingency tables, χ2 tests, correlations and multivariable logistic regression to explore relationships between variables (using SPSS v26 and STATA v16). We preselected four dependent variables critical to delivering care, presence or not of shortages of: PPE, staff, medicines, and other equipment (such as syringe drivers). Independent variables were: country/region, charity or public management, settings (comprising four settings), experiences with COVID-19 and level of busyness; criteria for inclusion in multiple regression analyses was p<0·10 in univariable analysis. We excluded variables exhibiting collinearity with independent variables already included if the variance inflation factor >10 or chi-square test, p < 0·05.

Free text comments were explored in Excel using content analysis^17^ to understand the impact of COVID-19 and the strategies, enablers and specific actions deployed by services.^17^

### Sample Size

The survey was intended to be largely descriptive yielding a breath of information where none is available. We aimed to have responses from >390 services, ∼130 inpatient palliative care units, 130 hospital palliative care teams, 130 home palliative care teams. Subgroups of this size (>105) are sufficient to detect differences with effect sizes of 0·35, using χ2 (p<0·05, df=5, power 80%).

## Results

In total, 489 questionnaire were commenced, 477 completed (completion rate 97·5%); of these 15 were duplicates and 2 triplicates of entries with the same name/email; 2 were invalid being from one researcher without a palliative care service, leaving 458 valid responses: 277 UK, 85 rest of Europe, 95 rest of the world, 1 missing country (Table 1; supplementary Table S1). Services were usually publicly (204, 46·4%), or charity managed (192, 43·6%); 19 (4·3%) were privately managed, 25 (5·7%) other; 18 missing. Charity managed services were less well integrated with national health services (mean (SD): 67·82 (19·75)) compared to publicly managed services (mean (SD): 75·10 (19·75)), (mean difference (95% CI): 7·28 (3·24-11·33); p < 0·001). Overall, 261 services provided inpatient palliative care units, 261 home care teams, 217 hospital palliative care teams, and 119 home nursing teams. Many services offered care in more than one setting, with publicly managed services often providing hospital teams combined with home care or inpatient care, and charity managed services often providing inpatient units combined with home palliative care teams and/or home nursing (supplementary Table S2).

**Table 1.**
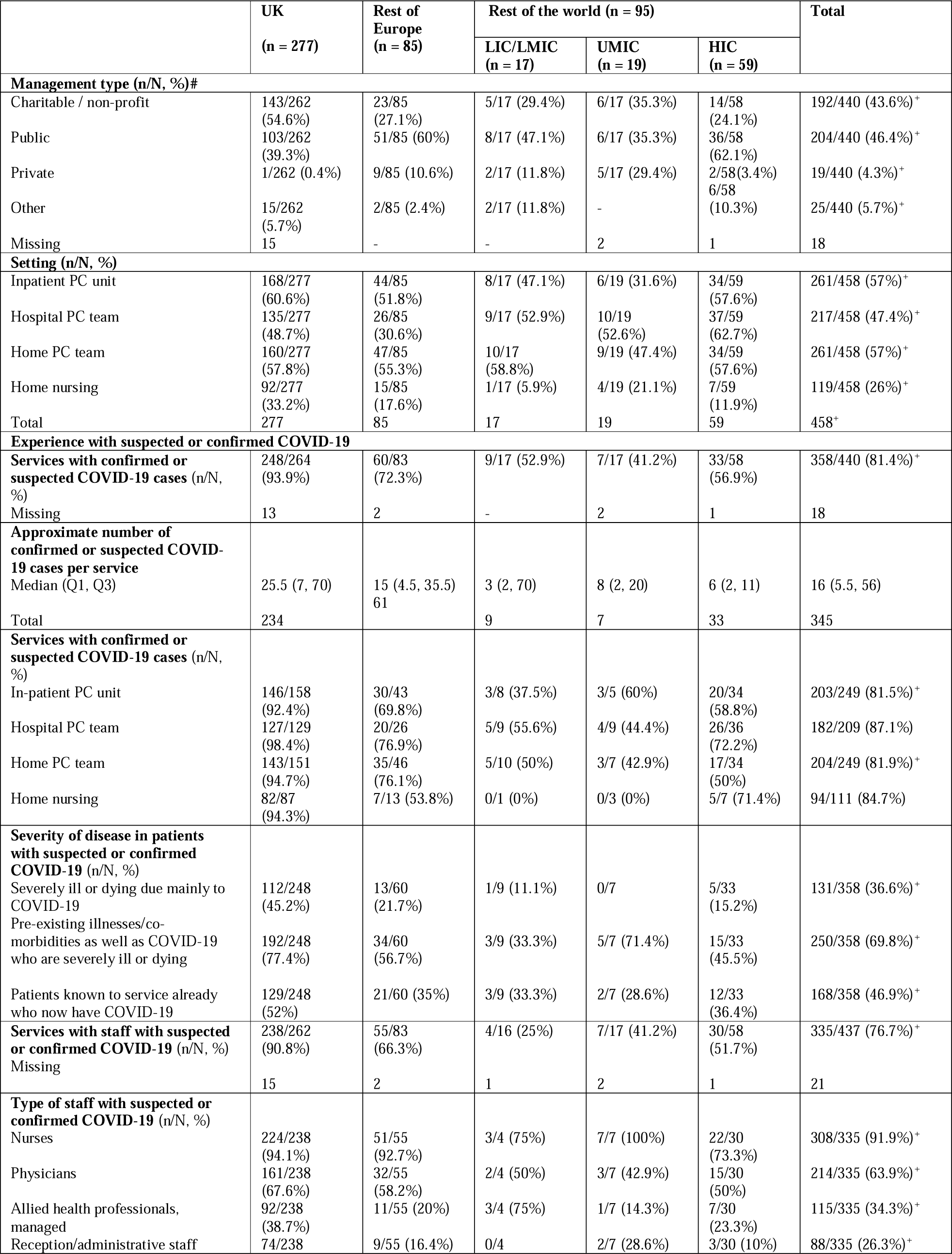
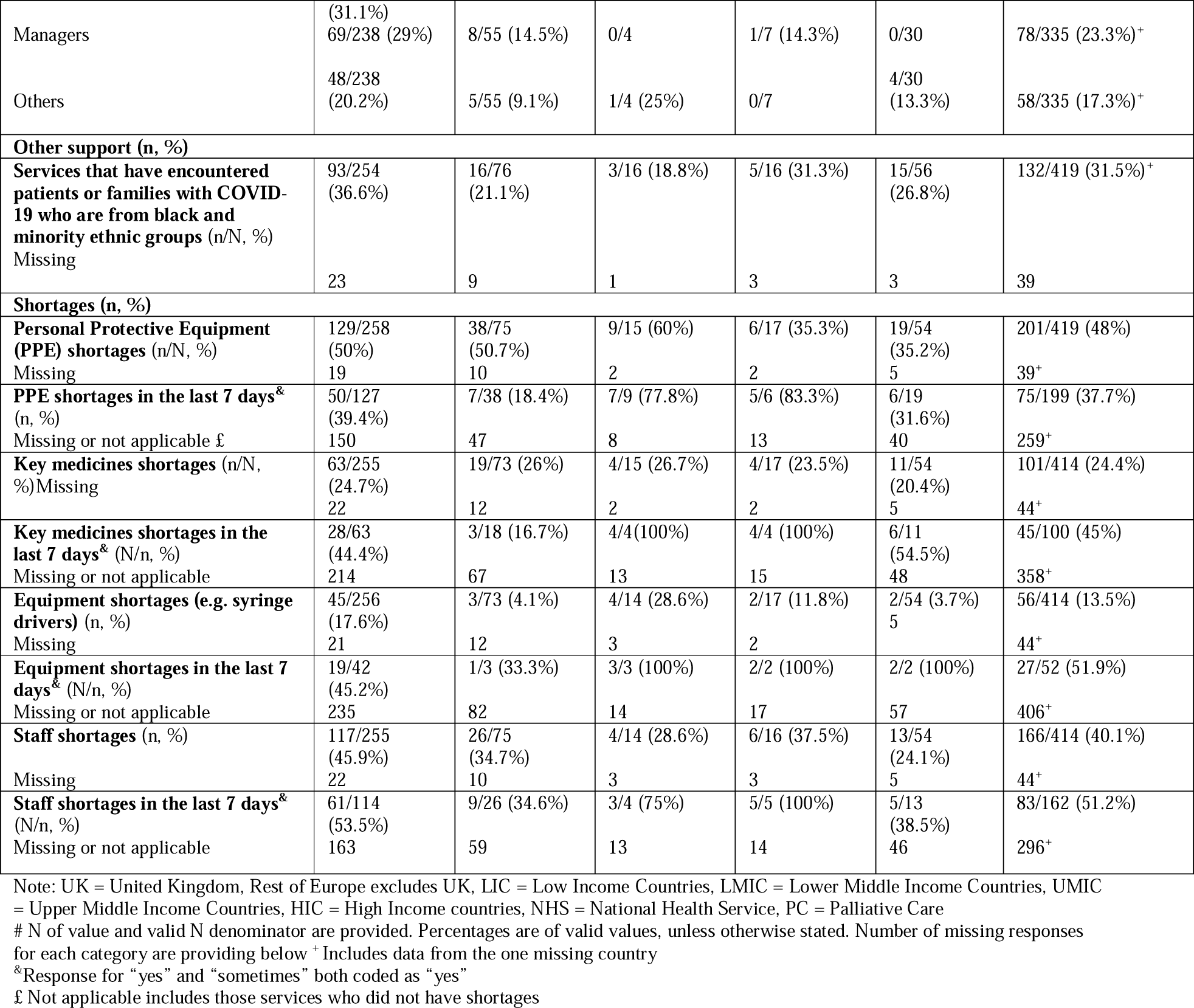
Characteristics of responding palliative care and hospice services by region

### Overall impact of COVID-19 on palliative and hospice services

Of all responding services, 91% changed how they worked as a result of COVID-19; 77% had staff who had suspected or confirmed cases of COVID-19; 29% had staff who were deployed elsewhere, and 36% had staff deployed to them. 81% of services had cared for patients with suspected or confirmed COVID-19, or both; of these, three main groups were cared for: patients with pre-existing illness or morbidities who were severely ill or dying from COVID-19 not previously known to palliative care (70% of services); patients dying from COVID-19 already known to the palliative care services (47%); and patients severely ill or dying from COVID-19 but without pre-existing illness or morbidities (37%) (Table 1, supplementary Table S1).

### Activities and changes in services

There was a pattern of reduced inpatient palliative care unit activity for free standing units, as patients who did not have COVID-19 did not want to be admitted to an inpatient unit for fear of contracting the infection. There was increased activity for home and hospital palliative care teams. Assistance from volunteers plummeted; of responding services who used volunteers, 79% used them much less, 10% slightly less. Many volunteers were from older age groups and therefore high risk. Other activities and changes included (supplementary tables S1, S3):

- a surge in number of patients cared for many times previously experienced pre-COVID-19;
- developing guidelines and education materials as none existed nationally;
- training and supporting other health professionals because the service could not support everyone in need;
- increased virtual and telephone monitoring (84 % of services increased this type of support for patients, and 95% of services in supporting other professionals);
- directly supporting continuous positive airway pressure or other ventilatory withdrawal;
- directly delivering or supporting community or district nursing services;
- reduction in support from volunteers, many of whom were shielding or not able to provide visits as previously; and
- switching from pro-active to reactive care because of the demand on services.

Services described how staff were often stressed by concerns for the patient’s health and their own, being unable to visit patients and adapting to new practices. They were sometimes frustrated by how their time and energy were consumed trying to source equipment, including PPE; without which staff got ill or could not deliver care (Figure 1). The lack of timely PPE and the ethical challenges this presented, in terms of whom they could visit, made them fearful and anxious. There were financial considerations, with some staff especially from charities, concerned for the viability of services (Figure 1; supplementary table S3).

**Figure 1.**
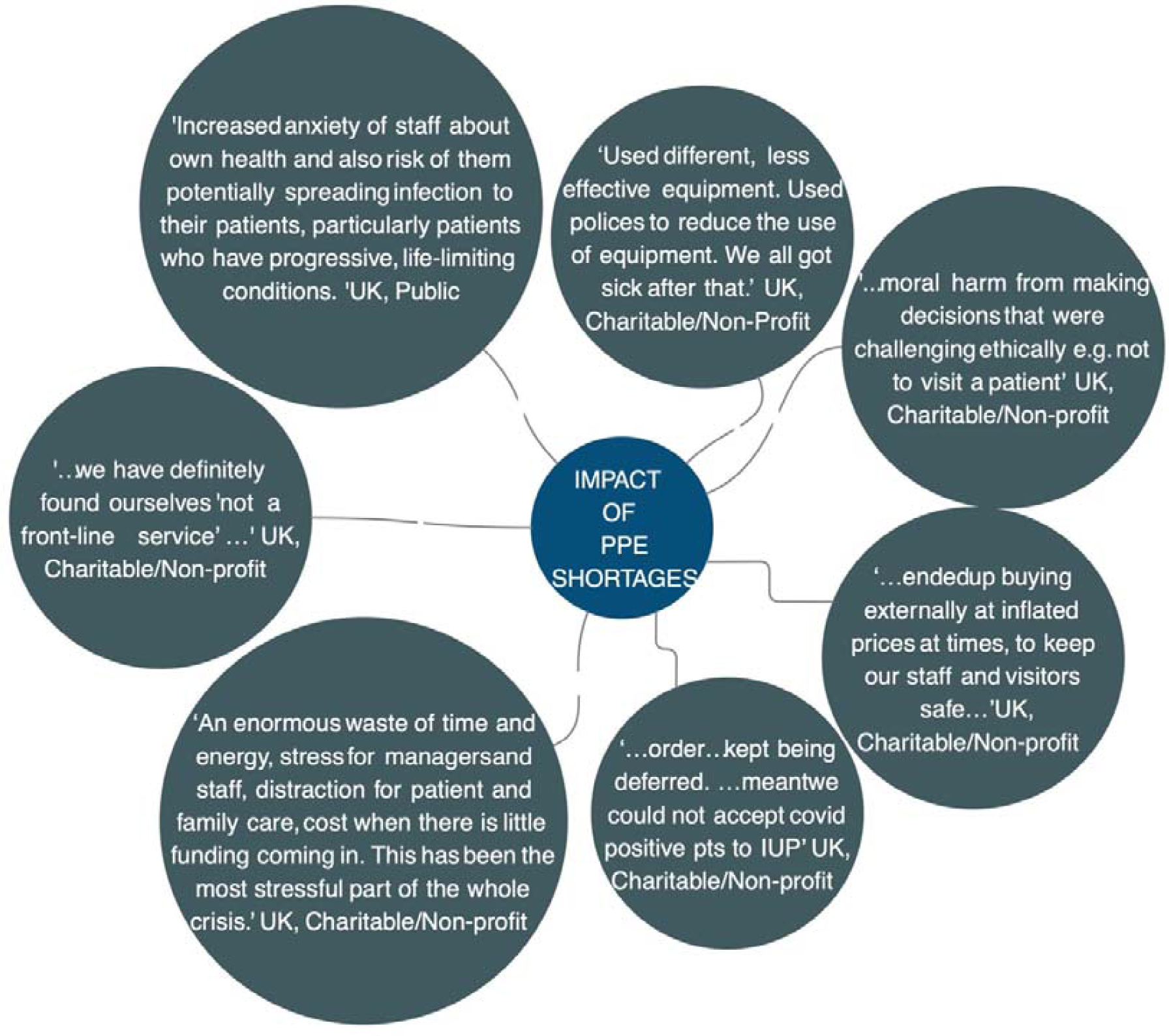
Quotes on the impact of PPE shortages on palliative care services.

### Shortages and associated factors

Of responding services, overall 48% reported shortages of PPE, 40% shortages of staff, 24% shortages of medicines, and 14% shortages of other equipment, commonly battery operated syringe drivers (Table 1).

#### PPE shortages

Being charity managed was associated with greater likelihood of PPE shortages compared with publicly managed (OR 3·07, 95% CI 1·81-5·20; p < 0.001) (Table 2, Figure 2). Inpatient palliative care units were more likely to have PPE shortages compared to other settings (OR 2·34, 95% CI 1·46-3·75; p < 0.001). Note that home nursing had high shortages of PPE (70%, figure 2) compared with other settings. Home nursing was most commonly provided by services also providing inpatient palliative care (see supplementary table S2); this may be confounding the relationship between inpatient palliative care units and PPE shortages. Hospital palliative care teams were less likely to have PPE shortages compared to other settings (OR 0·51, 95% CI 0·32-0·82; p = 0.005) The UK experienced similar levels of PPE shortages compared with elsewhere in Europe, but during the 7 days before completion of the survey, shortages were more common in the UK than in Europe (Table 1, Figure 2). Shortages were reported most commonly for masks (filtering facepiece, FFP2, FFP3), FIT testing kits for FFP3 masks, hospital scrubs, aprons, gloves, face shields, long sleeve gowns, hand gels, goggles and eye protection.

**Table 2.** Multivariate logistic regression: factors associated with shortages

| **Multivariate logistic regression: factors associated PPE shortages** |  |  |  |  |
| --- | --- | --- | --- | --- |
| **Variables** | **Odds ratio** | **CI<sub>lower</sub>** | **CI<sub>upper</sub>** | **P value** |
| **Unit management type** |  |  |  |  |
| Public | Ref |  |  |  |
| Charitable | 3.07 | 1.81 | 5.20 | P < 0.001 |
| Other | 0.97 | 0.46 | 2.05 | P = 0.95 |
| **Inpatient palliative care unit** |  |  |  |  |
| No | Ref |  |  |  |
| Yes | 2.34 | 1.46 | 3.75 | P < 0.001 |
| **Hospital palliative care team** |  |  |  |  |
| No | Ref |  |  |  |
| Yes | 0.51 | 0.32 | 0.82 | P = 0.005 |
| **Multivariate logistic regression: factors associated with staff shortages** |  |  |  |  |
| **Variables** | **Odds ratio** | **CI<sub>lower</sub>** | **CI<sub>upper</sub>** | **P value** |
| **Country** |  |  |  |  |
| UK | Ref |  |  |  |
| Rest of Europe | 0.63 | 0.37 | 1.07 | 0.09 |
| Rest of the world | 0.44 | 0.26 | 0.76 | 0.003 |
| **Multivariate logistic regression: factors associated with other equipment shortages** |  |  |  |  |
| **Variables** | **Odds ratio** | **CI<sub>lower</sub>** | **CI<sub>upper</sub>** | **P value** |
| **Inpatient palliative care unit** |  |  |  |  |
| No | Ref |  |  |  |
| Yes | 0.35 | 0.18 | 0.65 | 0.001 |
| **Country** |  |  |  |  |
| UK | Ref |  |  |  |
| Rest of Europe | 0.15 | 0.04 | 0.51 | 0.002 |
| Rest of the world | 0.46 | 0.20 | 1.08 | 0.07 |
| **Level of busyness** |  |  |  |  |
| About the same | Ref |  |  |  |
| A lot more busy | 10.81 | 3.10 | 37.71 | <0.001 |
| Slightly more busy | 5.41 | 1.48 | 19.76 | 0.01 |
| Slightly less busy | 3.24 | 0.81 | 12.89 | 0.10 |
| Much less busy | 1.32 | 0.21 | 8.34 | 0.77 |

Note: No multivariate logistic regression analysis could be carried out for medicines shortages because only one variable (inpatient hospice/palliative care unit) was relevant from the univariable analysis
REF: Reference category

**Figure 2.**
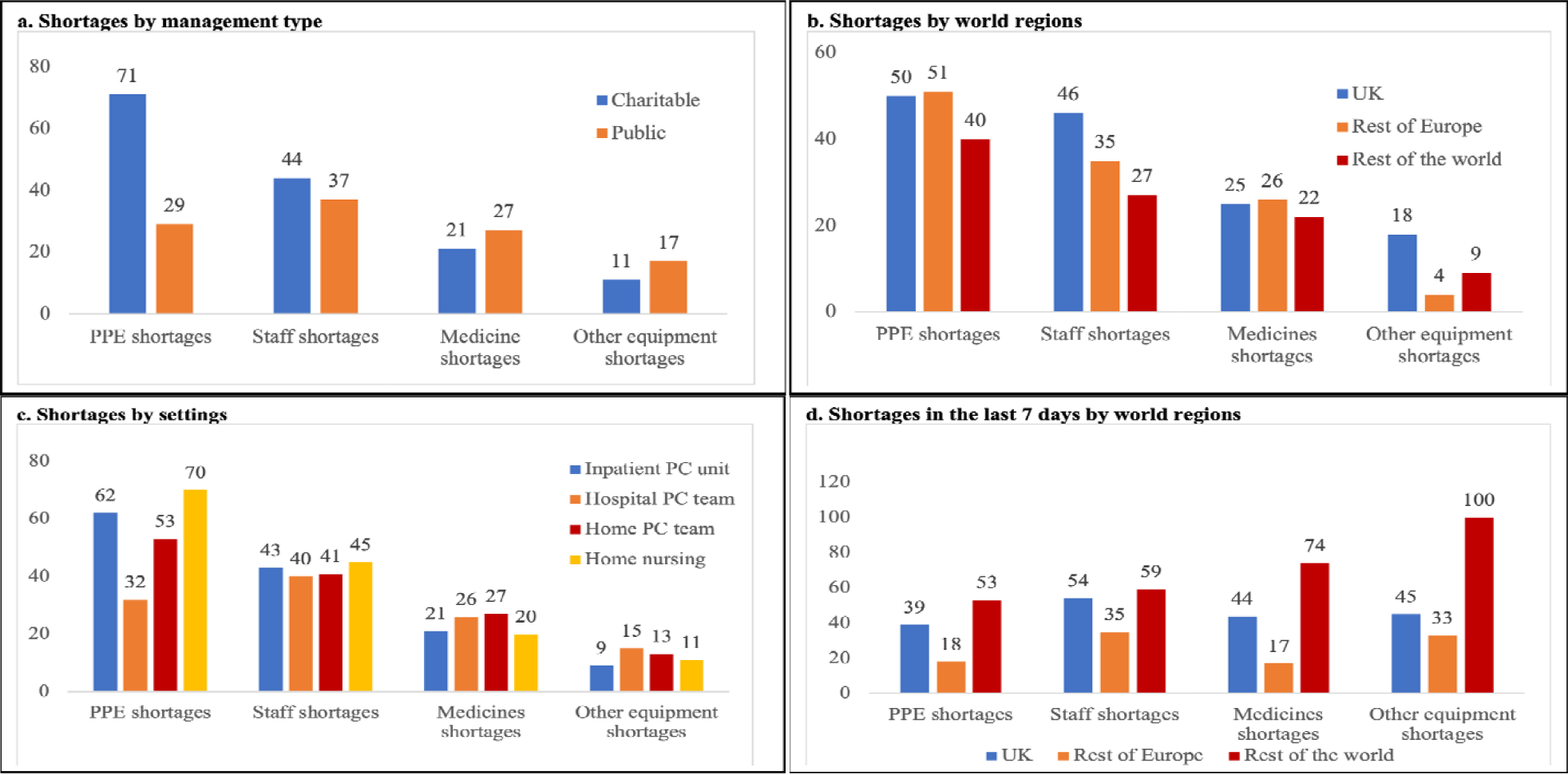
Percentage of shortages by management type, world regions and settings.

#### Staff shortages

Staff shortages were higher in the UK (46%) than other countries (27%) (Figure 2); compared with the UK, the rest of the world had lower odds of staff shortages (regression analysis OR 0·44, 95% CI 0·26-0·76; p = 0.003). Other factors were not significantly associated with staff shortages in regression analysis. Services reported shortages in specialist palliative care teams, of doctors (consultants, specialty doctors, middle grade and junior doctors), nurses (advanced practitioners, clinical nurse specialists, community nurses, community palliative care nurses, registered general nurses, ward manager), allied health professionals (healthcare assistants, occupational therapists, pharmacists, pharmacy assistants, physiotherapists, social workers), administrative and housekeeping staff.

**Figure 2.**
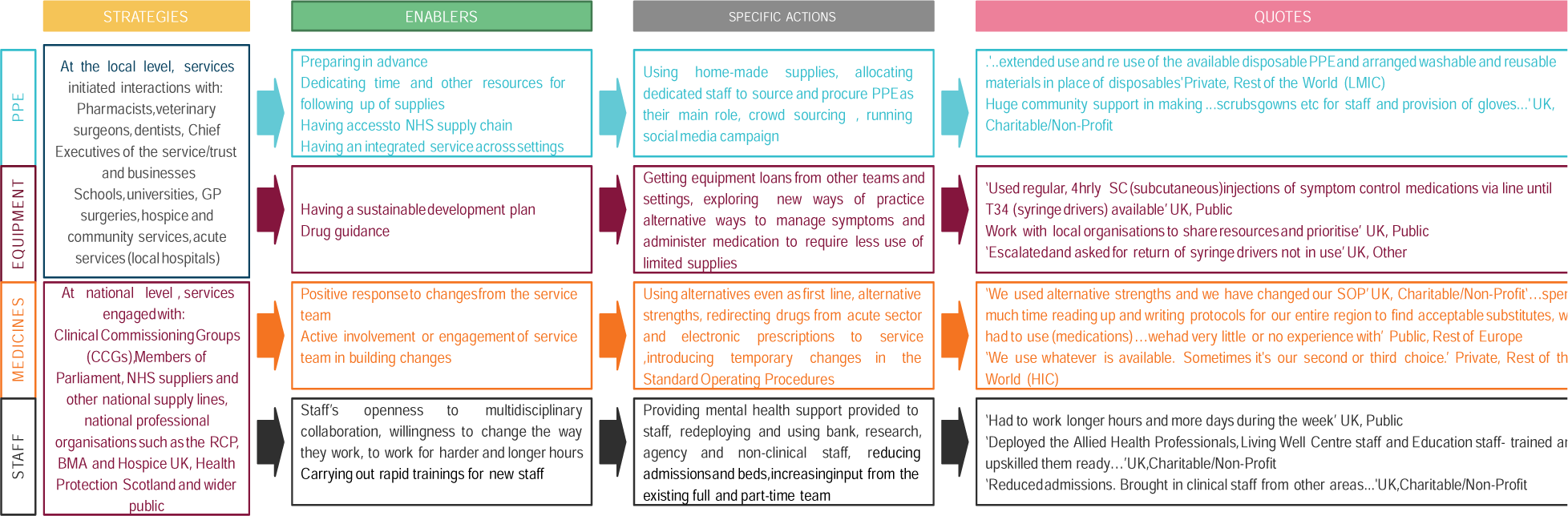
Strategies, enablers, specific actions and quotes about shortages.

#### Medicines shortages

The commonly reported top three medicine shortages were: levomepromazine, midazolam (used for symptoms of agitation and delirium) and alfentanil (used for pain and breathlessness, commonly used in the UK in severe renal impairment, when morphine is contra-indicated). Shortages of medicines affected 20-27% of settings (Figure 2), and were similar across regions. Only one variable (inpatient palliative care unit) met our criteria for inclusion from the univariable analysis (at P=0.07), so multiple regression analysis was not attempted.

#### Other equipment shortages

Services reported shortages or delays in access to other equipment, especially syringe drivers/pumps, syringe pump lines, butterfly needles and tympanic thermometer covers. Inpatient palliative care units were less likely to have other equipment shortages compared to other settings (OR 0·35, 95% CI 0·18-0·65; p = 0.001) (Figure 2). When compared to the UK, being elsewhere in Europe was associated with decreased odds of other equipment shortages (OR 0·15, 0·04-0·51; p = 0.002). Services that reported being a lot more busy (OR 10·81, 95% CI 3·10-37·71; p < 0.001) and slightly more busy (OR 5·41, 95% CI 1·48-19·76; p = 0.01) were more likely to have other equipment shortages than other services.

### Response to shortages

Services expended huge efforts to procure PPE, some deployed staff to find PPE as their main role. Services contacted local vets, schools, dentists, universities, hospitals, businesses, health services, and national supply lines, government, professional organisations and the wider public. Some made their own supplies, crowd sourced and ran social media campaigns (Figure 3).

In response to staff shortages, multidisciplinary full and part-time staff worked longer hours, extra shifts and flexibly collaborated. Services reduced hospice beds; day-care services were closed, staff deployed to the community. Bank, research, agency and non-clinical staff were used. New staff were rapidly trained. Some services drafted emergency staffing plans. Mental health support was provided. As one service lead summarised ‘the impact of the multiple ‘threats’ to self [that staff felt] should not be underestimated’ as the staff are worried about themselves, their families, their colleagues, their patients, working in unfamiliar areas and experiencing ‘…[Acute Respiratory Disease Syndrome] as a distressing mode of death’.

In response to medicine shortages, services contacted local services and pharmacists (Figure 3). Some used alternative medicines even as first line, alternative strengths (lower or higher concentrations), or reduced the numbers of vials when prescribing home medications. For example, for shortages of levomepromazine, services prioritised it for agitation, using alternatives for controlling nausea and vomiting. Services introduced temporary changes in Standard Operating Procedures (SOP) and developed new SOPs on drugs reuse. The positive response and active involvement of local teams enabled the adaptation, as one service reported ‘…staff [were] very engaged and responsive to our requests for frequent change of practice.’

In response to shortages of other equipment, services tried to get loans from other teams and settings (Figure 3). The teams explored alternative ways to manage symptoms and administer medication to require less use of limited equipment. For example, faced with a shortage of syringe drivers (small, portable, battery powered infusion pumps suitable for use in hospital, care homes and at home to give medicines for breathlessness, pain and agitation), services carried out risk assessments, programmed alternative pumps, such as 50 ml infusion devices, gave 4 hourly subcutaneous injections, or considered using transdermal patches rather than subcutaneous infusions.

## Discussion

We report the first multinational survey on the response of and challenges to palliative care services during the COVID-19 pandemic. Across all settings palliative care services rapidly adjusted and increased the volume and type of provision, caring for people with COVID-19, as well as for existing patients. Patients dying from and with severe symptoms due to COVID-19 were in three main categories: patients with underlying conditions and/or multimorbid disease not previously known to palliative care, patients already known to palliative care services, and patients previously healthy, who were now dying from COVID-19. Palliative care became very busy, especially in areas with high COVID-19 prevalence. Care shifted to community and hospital support from inpatient palliative care units. Staff were also infected by COVID-19. Services experienced multiple shortages of equipment, including PPE, staff and medicines that limited their ability to respond. Our two prior null hypotheses were rejected. There were differences in shortages between services with different management type and between settings.

Palliative care staff responded dynamically; they provided care directly to patients and families across hospitals and the community, supported other clinical staff through training, symptom management, communication and care guidance, supported decision making and filled gaps in care. They used their expertise to propose strategies to deal with medicines shortages. These contributions have been vital; clinicians with limited or no palliative care experience had to provide end of life care to patients and to support their families.^18^ Palliative care clinicians adapted and innovated quickly, possibly helped by prior experience with patients with different diseases and with multimorbidity. Palliative care puts the person before their disease wherever they are cared for: it is neither disease nor setting specific. The symptoms and problems of severe COVID-19 are commonly breathlessness and agitation,^19,20^ both familiar to palliative care clinicians. Current case reports suggest that these symptoms in end stage COVID-19 can be alleviated with low doses of opioids and benzodiazepines, delivered subcutaneously with a battery operated syringe driver.^19,20^ Palliative care has expertise in holistic end of life care, care for older people and those with multimorbidity.^6^ This flexibility, expertise and learning will be crucial to the international response to COVID-19, especially as cases of COVID-19 continue to rise across the globe.

Our study identified three different groups of patients, and these may require different approaches. A parallel planning approach may be needed for patients with uncertain trajectories, as is often used among patients with other uncertain prognosis in serious illness, such as haematological cancers, and in children’s intensive/palliative care.^21,22^ Parallel planning provides for two sets of plans, run side by side. Both plans aim to ensure symptom management and the best in care: one plan is towards improvement or recovery, the second plan is made in case the patient deteriorates or begins to die.^22^

Such parallel plans may be important in alleviating concerns about care rationing and communication, which have been identified in public consultation.^23^ In depth consideration of the changes and challenges encountered in advance care planning during COVID-19 would shed light on how to improve communication and care planning, and support individual patient and family values at this critical time.

Despite efforts to respond and their resilience, palliative care services experienced considerable shortages of PPE, staff, medicines and other equipment. PPE shortages especially affected charity managed services. Almost half of services were charity managed; these had lower levels of integration with national health systems than publicly managed services. It is six years since the World Health Assembly resolution in 2014, calling for better integration for palliative care into health care systems; a declaration endorsed by all countries.^24^ Our findings reveal little progress and highlight the consequences. Most countries of the globe lack palliative care doctors, nurses, allied health professionals and trainees to meet their needs.^25,26^ Undergraduate doctors and nurses receive scant training in palliative care, leaving them lacking in confidence and skills in symptom management, communication and care, and resulting in stress following complex encounters.^27,28^ Undergraduate and postgraduate training from expert palliative care clinicians is urgent, as well as developing an adequate palliative care workforce to support services and patients day to day, especially as COVID-19 cases increase.

Home nursing and inpatient palliative care units were most seriously affected by PPE shortages. This continued even in the 7 days before survey completion, especially for home nursing services and in the UK. Many palliative care patients chose not to come to hospital, preferring care at home for fear of contracting COVID-19 and/or wishing to remain close to those important to them, a result supported by research in other pandemics,^13,29^ and data from the first wave of the COVID-19 pandemic.^30^ Taken together these findings are concerning. If patients are to remain in the community, then the community and care homes, as well as hospitals, need sufficient resources to be able to provide care; this includes protective equipment, syringe drivers to deliver subcutaneous infusions to control symptoms and sufficient staff and medicines. Legislation on the reuse of medicines in the community, especially at times of shortages, may need to be revised. The role of free standing inpatient palliative care units during pandemics is worthy of consideration and planning: could their staff proactively be diverted to the community (as occurred in many settings in our study), could they be diverted to hospital palliative care teams or care homes (both settings needing additional support), or could they provide an alternative or rehabilitation/step down care from hospitals? Any option would need planning, training, and possibly a different skill mix.

In conclusion, palliative care services responded actively but felt ignored by many national health systems during the COVID-19 pandemic, despite supporting patients who were dying or had severe symptoms, supporting their families/carers and supporting other professionals to deliver care. Services provided expertise in symptom management and holistic care while facing shortages of equipment, staff and medicines. The crucial role of palliative care during pandemics must be better recognised and integrated. This is particularly the case for charity managed services and those providing care in people’s homes. Beyond COVID-19, this research has shed light on the limited integration of palliative care, the urgent need to increase its workforce and a need for palliative skills to be a core part of the training of clinicians, including specialists in infectious diseases.

## Supporting information

Supplementary Files with Appendices I II and III

CHERRIES, Checklist of MORECare Statement and STROBE Checklist

## Data Availability

Applications for use of the survey data can be made for up to 10 years, and will be considered on a case by case basis on receipt of a methodological sound proposal to achieve aims in line with the original protocol. The study protocol is available on request. All requests for data access should be
addressed to the Chief Investigator via the details on the CovPall website
(https://www.kcl.ac.uk/cicelysaunders/research/evaluating/covpall-study, and) and will be reviewed by the Study Steering Group.

## Contributions and Acknowledgements

### Contributors

IJH is the grant holder and chief investigator; KES, MM, FEM, CW, NP, LKF, SB, MBH and AOO were co-applicants for funding. IJH and CW with critical input from all authors wrote the protocol for the CovPall study. MBH, AOO, RC and LD co-ordinated data collection and liaised with centres, with input from IJH, FEM, CW, NP and LKF. AOO, MBH, IJH and LF analysed the data. All authors had access to all study data, discussed the interpretation of findings and take responsibility for data integrity and analysis. AOO, IJH, MBH drafted the manuscript. AOO and MBH are equal contributing first authors. All authors contributed to the analysis plan and provided critical revision of the manuscript for important intellectual content. IJH is the guarantor, senior author and author for correspondence.

### CovPall Study Team

Professor Irene J Higginson (Chief Investigator), Dr Sabrina Bajwah (Co-I), Dr Matthew Maddocks (Co-I), Professor Fliss Murtagh (Co-I), Professor Nancy Preston (Co-I), Dr Katherine E Sleeman (Co-I), Professor Catherine Walshe (Co-I), Professor Lorna K Fraser (Co-I), Dr Mevhibe B Hocaoglu (Co-I), Dr Adejoke O Oluyase (Co-I), Dr Andrew Bradshaw, Lesley Dunleavy and Rachel L Cripps.

#### CovPall Study Partners

Hospice UK, Marie Curie, Sue Ryder, Palliative Outcome Scale Team, European Association of Palliative Care (EAPC), Together for Short Lives and Scottish Partnership for Palliative Care.

## Acknowledgments

This study was part of CovPall, a multi-national study, supported by the Medical Research Council, National Institute for Health Research Applied Research Collaboration South London and Cicely Saunders International. We thank all collaborators and advisors. We thank all participants, partners, PPI members and our Study Steering Group. We gratefully acknowledge technical assistance from the Precision Health Informatics Data Lab group (https://phidatalab.org) at National Institute for Health Research (NIHR) Biomedical Research Centre at South London and Maudsley NHS Foundation Trust and King’s College London for the use of REDCap for data capture.

## Data sharing

Applications for use of the survey data can be made for up to 10 years, and will be considered on a case by case basis on receipt of a methodological sound proposal to achieve aims in line with the original protocol. The study protocol is available on request. All requests for data access should be addressed to the Chief Investigator via the details on the CovPall website (https://www.kcl.ac.uk/cicelysaunders/research/evaluating/covpall-study, and) and will be reviewed by the Study Steering Group.

