## Supplementary Files with Appendices I II and III for "The challenges of caring for people dying from COVID-19: a multinational, observational study of palliative and hospice services (CovPall)"

#### Supplementary Material

Appendix I

CovPall

##### Box S 1 Procedures for CovPall survey

Services were identified and contacted through national and multinational gatekeeper palliative care and hospice organisations (see acknowledgements and supplementary appendix III for details) and asked that their medical or nursing lead, or their nominee, complete an on-line survey available via a link. The email attached the participant information sheet with details of the study rationale, ethical approval, data protection and management, investigators and contacts for further information or concern. No incentives were offered for completion. Completion was taken to indicate informed consent. The CovPall study was presented at relevant on-line meetings and discussed with gatekeeper organisations and in blogs, to inform the methods, questions and to raise awareness and engagement.

We developed and piloted a secure, password-protected web-based data entry portal (in the Research Electronic Data Capture (REDCap) at the study coordinating centre, King's College London. Services could keep their identity hidden if they wished, but most chose to provide an email for contact. Data were anonymised before analysis.

The questionnaire was developed and piloted by the CovPall study team building on an earlier survey of Italian hospices, adding questions on the impact of and response to COVID-19. It was intended to be brief, taking around 30 minutes to complete as we recognised staff were busy, and comprised seven sections: region and responder; Palliative Care (PC)/hospice management and services offered pre-COVID-19; experiences of patients and staff with COVID-19; changes to services overall; changes in specific settings; shortages experienced; symptom management. Free-text explanatory comments were invited in all sections and respondents also indicated whether and how they could be contacted. Respondents could save and complete the questionnaire later if they wished. The questionnaire is available in supplementary appendix II.

The survey opened on April 23rd and closed July 31st 2020. The study coordinating centre replied actively to respondents queries or requests for help.

In most cases, data collection and entry was done electronically via the REDCap link directly by the services without problems, however a few palliative care services had difficulties accessing the online survey due to local computer procedures. In these instances they were sent the survey in paper format, completed this and returned it to King's to enter this data, or were interviewed over the phone and data entry sheet was completed by the researchers. The research team also audited the data weekly to ensure data entry completeness and sent monthly missing data and incomplete entry reports to the research associates and administrators, and where consent permitted to relevant respondents to check validity.

**Table S 1 Characteristics of responding palliative care and hospice services by region**

| | UK (n = 277) | Rest of Europe (n = 85) | Rest of the world (n = 95) | | | Total |
|---|---|---|---|---|---|---|
| | | | LIC/LMIC (n = 17) | UMIC (n = 19) | HIC (n = 59) | |
| **Role of respondents** (n/N, %)**#** | | | | | | |
| Medical director/ lead medical clinician | 97/274 (35.4%) | 42/85 (49.4%) | 11/17 (64.7%) | 7/19 (36.8%) | 26/58 (44.8%) | 183/453 (40.4%) |
| Nurse director/ lead nurse clinician | 69/274 (25.2%) | 8/85 (9.4%) | 2/17 (11.8%) | 2/19 (10.5%) | 7/58 (12.1%) | 88/453 (19.4%) |
| Other | 108/274 (39.4%) | 35/85 (41.2%) | 4/17 (23.5%) | 10/19 (52.6%) | 25/58 (43.1%) | 182/453 (40.2%) |
| Missing | 3 | - | - | - | 1 | 5[+] |
| **Management type** (n/N, %)**#** | | | | | | |
| Charitable / non-profit | 143/262 (54.6%) | 23/85 (27.1%) | 5/17 (29.4%) | 6/17 (35.3%) | 14/58 (24.1%) | 192/440 (43.6%)[*] |
| Public | 103/262 (39.3%) | 51/85 (60%) | 8/17 (47.1%) | 6/17 (35.3%) | 36/58 (62.1%) | 204/440 (46.4%)[+] |
| Private | 1/262 (0.4%) | 9/85 (10.6%) | 2/17 (11.8%) | 5/17 (29.4%) | 2/58(3.4%) | 19/440 (4.3%)[+] |
| Other | 15/262 (5.7%) | 2/85 (2.4%) | 2/17 (11.8%) | 0/17 (0%) | 6/58 (10.3%) | 25/440 (5.7%)[+] |
| Missing | 15 | - | - | 2 | 1 | 18 |
| **Type of service** (n/N, %) | | | | | | |
| Adult only | 247/277 (89.2%) | 68/85 (80%) | 7/17 (41.2%) | 9/19 (47.4%) | 40/59 (67.8%) | 371/458 (81%)[+] |
| Children only | 16/277 (5.8%) | 5/85 (5.9%) | 0/17 (0%) | 3/19 (15.8%) | 5/59 (8.5%) | 29/458 (6.3%)[+] |
| Both adult and children | 11/277 (4%) | 10/85 (11.8%) | 9/17 (52.9%) | 7/19 (36.8%) | 14/59 (23.7%) | 52/458 (11.4%)[*] |
| None indicated | 3/277 (1.1%) | 2/85 (2.4%) | 1/17 (5.9%) | 0/19 (0%) | 0/59 (0%) | 6/458 (1.3%)[+] |
| **Setting** (n/N, %) | | | | | | |
| Inpatient PC unit | 168/277 (60.6%) | 44/85 (51.8%) | 8/17 (47.1%) | 6/19 (31.6%) | 34/59 (57.6%) | 261/458 (57%)[*] |
| Hospital PC team | 135/277 (48.7%) | 26/85 (30.6%) | 9/17 (52.9%) | 10/19 (52.6%) | 37/59 (62.7%) | 217/458 (47.4%)[+] |
| Home PC team | 160/277 (57.8%) | 47/85 (55.3%) | 10/17 (58.8%) | 9/19 (47.4%) | 34/59 (57.6%) | 261/458 (57%)[*] |
| Home nursing | 92/277 (33.2%) | 15/85 (17.6%) | 1/17 (5.9%) | 4/19 (21.1%) | 7/59 (11.9%) | 119/458 (26%)[+] |
| Total | 277 | 85 | 17 | 19 | 59 | 458[+] |
| **Setting** (n/N, %) | | | | | | |
| None indicated | 3/277 (1.1%) | 1/85 (1.2%) | 1/17 (5.9%) | 0/19 (0%) | 0/59 (0%) | 5/458 (1.1%)[+] |
| Single setting | 110/277 (39.7%) | 49/85 (57.6%) | 8/17 (47.1%) | 11/19 (57.9%) | 24/59 (40.7%) | 202/458 (44.1%)[+] |
| Multiple settings (2 or more settings | 164/277 (59.2%) | 35/85 (41.2%) | 8/17 (47.1) | 8/19 (42.1%) | 35/59 (59.3%) | 251/458 (54.8%)[*] |
| **% funding usually from NHS** | | | | | | |
| Mean (SD) | 28.3 (13) | 41.4 (35) | 20 (34.6) | 24.6 (42.6) | 34.1 (35.3) | 30.1 (20.9) |
| Median (IQR) | 25 (20, 35.5) | 31.5 (9.8, 70) | 0 (0, 50) | 10 (0, 56.5) | 40 (0, 60) | 25 (19, 40) |
| Total | 137 | 22 | 5 | 5 | 11 | 181[+] |
| Missing | 140 | 63 | 12 | 14 | 48 | 277 |
| **Level of Integration with NHS** | | | | | | |
| Mean (SD) | 74 (16.8) | 72.1 (22.7) | 55.4 (31.8) | 51.9 (27.5) | 68.3 (25.3) | 71.5 (21) |
| Median (IQR) | 76 (63.8, 87) | 76.5 (66.3, 87) | 59 (28.3, 78.5) | 49 (29, 77) | 73 (49.8, 90) | 75 (60.8, 87) |
| Total | 250 | 80 | 16 | 15 | 52 | 414[+] |
| Missing | 27 | 5 | 1 | 4 | 7 | 44 |
| **Information about services offered before COVID-19 pandemic** | | | | | | |
| **Bereavement services** (n/N, %)**#** | 201/265 (75.8%) | 58/83 (69.9%) | 13/17 (76.5%) | 12/17 (70.6%) | 42/58 (72.4%) | 327/441 (74.1%)[*] |
| Missing | 12 | 2 | - | 2 | 1 | 17 |
| **Bereavement services offered only to families/friends of patients who had been cared for by your service** (n/N, %)**#** | 135/199 (67.8%) | 49/58 (84.5%) | 11/13 (84.6%) | 9/12 (75%) | 26/42 (61.9%) | 231/325 (71.1%)[*] |
| Missing | 78 | 27 | 4 | 7 | 17 | 133 |

| | | | | | | |
|---|---|---|---|---|---|---|
| **Use of a risk assessment tool to help you decide how to target bereavement services** (n/N, %)**#**<br>Missing | 82/189 (43.4%)<br>88 | 10/57 (17.5%)<br>28 | 4/13 (30.8%)<br>4 | 6/12 (50%)<br>7 | 13/42 (31%)<br>17 | 115/314 (36.6%)+<br>144 |
| **Volunteer roles within service**<br>Direct patient/family facing support (e.g. befriending, home visits, in-patient unit care, family support groups/visiting etc) (n/N, %) | 166/277 (59.9%) | 44/85 (51.8%) | 13/17 (76.5%) | 15/19 (78.9%) | 30/59 (50.8%) | 269/458 (58.7%)* |
| **Volunteer roles within service**<br>Indirect patient/family facing support (e.g. reception functions, refreshments, driving/transport etc) (n/N, %) | 164/277 (59.2%) | 31/85 (36.5%) | 5/17 (29.4%) | 7/19 (36.8%) | 30/59 (50.8%) | 238/458 (52%)* |
| **Volunteer roles within service**<br>Back office functions (e.g. finance support, maintenance, gardening etc) (n/N, %) | 162/277 (58.5%) | 28/85 (32.9%) | 5/17 (29.4%) | 5/19 (26.3%) | 21/59 (35.6%) | 222/458 (48.5%)* |
| **Volunteer roles within service**<br>Fundraising functions (e.g. shop volunteers, lottery etc) (n/N, %) | 153/277 (55.2%) | 26/85 (30.6%) | 3/17 (17.6%) | 4/19 (21.1%) | 22/59 (37.3%) | 209/458 (45.6%)* |
| **Volunteer roles within service**<br>Other (n/N, %) | 34/277 (12.3%) | 5/85 (5.9%) | 0/17 (0%) | 3/19 (15.8%) | 9/59 (15.3%) | 51/458 (11.1%)+ |
| **Use of remote consultations to help support patient care or education before COVID-19**<br>Telephone support for education (n/N, %) | 56/277 (20.2%) | 29/85 (34.1%) | 7/17 (41.2%) | 9/19 (47.4%) | 24/59 (40.7%) | 125/458 (27.3%)+ |
| **Use of remote consultations to help support patient care or education before COVID-19**<br>Telephone support for clinical care (n/N, %) | 179/277 (64.6%) | 54/85 (63.5%) | 13/17 (76.5%) | 11/19 (57.9%) | 45/59 (76.3%) | 303/458 (66.2%)* |
| **Use of remote consultations to help support patient care or education before COVID-19**<br>Telehealth/video support/e-learning for education (n/N, %) | 88/277 (31.8%) | 11/85 (12.9%) | 1/17 (5.9%) | 6/19 (31.6%) | 21/59 (35.6%) | 127/458 (27.7%)+ |
| **Use of remote consultations to help support patient care or education before COVID-19**<br>Telehealth/ video support/e-learning for clinical care (n/N, %) | 54/277 (19.5%) | 11/85 (12.9%) | 4/17 (23.5%) | 4/19 (21.1%) | 26/59 (44.1%) | 99/458 (21.6%)+ |
| **Number of beds in in-patient hospice/palliative care unit**<br>Mean (SD)<br>Median (IQR)<br>Total<br>Missing | 16.3 (9.3)<br>14.5 (10, 20)<br>166<br>111 | 20.5 (31.5)<br>10.5 (8, 19.5)<br>44<br>41 | 138.4 (348.5)<br>11 (6.3, 42.5)<br>8<br>9 | 20.7 (17.7)<br>14.5 (12.3, 27.5)<br>6<br>13 | 45.5 (171.8)<br>11 (8, 20.5)<br>33<br>26 | 24.6 (87.8)<br>14 (10, 20)<br>258+<br>200 |
| **Approximate number of new patients seen annually in in-patient palliative care unit**<br>Mean (SD)<br>Median (IQR)<br>Total<br>Missing | 348.5 (292.1)<br>300 (200, 400)<br>146<br>131 | 265.6 (257.1)<br>200 (112.5, 328)<br>44<br>41 | 2756.9 (6970)<br>395 (87.5, 500)<br>8<br>9 | 185.5 (288.8)<br>50 (38.7, 400)<br>5<br>14 | 245.6 (197.9)<br>200 (80, 350)<br>31<br>28 | 397.8 (1313)<br>285 (177, 400)<br>235+<br>223 |
| **Approximate number of new patients seen annually by hospital palliative care team**<br>Mean (SD)<br>Median (IQR)<br>Total<br>Missing | 1131 (950.4)<br>950 (470, 1677.5)<br>120<br>157 | 504.4 (419.9)<br>475 (97.5, 812.5)<br>26<br>59 | 2228 (3884)<br>300 (31, 4455)<br>9<br>8 | 268.5 (292)<br>165 (72.5, 425)<br>10<br>9 | 784.1 (2003.6)<br>300 (100, 700)<br>35<br>24 | 995.1 (1415.7)<br>700 (200, 1200)<br>200<br>258+ |
| **Hospital palliative care team supported** (n/N, %) | | | | | | |

| | | | | | | |
|---|---|---|---|---|---|---|
| Acute hospital<br><br>Community hospital | 127/135 (94.1%)<br>34/135 (25.2%) | 26/26 (100%)<br>8/26 (30.8%) | 7/9 (77.8%)<br><br>4/9 (44.4%) | 9/10 (90%)<br><br>3/10 (30%) | 30/37 (81.1%)<br>15/37 (40.5%) | 199/217 (91.7%)<br><br>64/217 (29.5%) |
| **Hospital palliative care team offered 24 hour support for patients** (n/N, %) | 101/135 (74.8%) | 12/26 (46.2%) | 7/9 (77.8%) | 8/10 (80%) | 28/37 (75.7%) | 156/217 (71.9%) |
| **Approximate number of new patients seen annually by home palliative care team**<br>Mean (SD)<br><br>Median (IQR)<br><br>Total<br>Missing | <br><br><br>947.5 (756.9)<br>775 (363, 1400)<br>139<br>138 | <br><br><br>454 (576.6)<br><br>300 (100, 550)<br>47<br>38 | <br><br><br>748 (1618.4)<br><br>100 (50, 612.5)<br>9<br>8 | <br><br><br>456.3 (573.5)<br>175 (62.5, 750)<br>8<br>11 | <br><br><br>489.4 (949.4)<br>175 (63.8, 400)<br>32<br>27 | <br><br><br>761.6 (819.8)<br><br>545 (170, 1062.3)<br><br>236+<br>222 |
| **Home palliative care team offered support to patients in care homes** (n/N, %) | 138/148 (93.2%) | 42/47 (89.4%) | 6/10 (60%) | 7/8 (87.5%) | 32/33 (97%) | 226/247 (91.5%)* |
| **Home palliative care team offered 24 hour support for patients** (n/N, %) | 103/160 (64.4%) | 32/47 (68.1%) | 7/10 (70%) | 9/9 (100%) | 23/34 (67.6%) | 174/261 (66.7%)+ |
| **Approximate number of new patients seen annually by home care**<br>Mean (SD)<br><br>Median (IQR)<br><br>Total<br>Missing | <br><br><br>430.9 (555.3)<br>300 (164, 436.5)<br>73<br>204 | <br><br><br>99.3 (122.6)<br><br>50 (30, 120)<br><br>15<br>70 | <br><br><br>90<br><br>90 (90, 90)<br><br>1<br>16 | <br><br><br>450 (312.2)<br><br>350 (200, -)<br><br>3<br>16 | <br><br><br>195.8 (129)<br><br>200 (55, 331.3)<br>6<br>53 | <br><br><br>362.8 (500.8)<br><br>250 (90, 381.3)<br><br>98<br>360+ |
| **Home nursing service offered 24 hour support for patients** (n/N, %) | 54/92 (58.7%) | 6/15 (40%) | 0/1 (0%) | 3/4 (75%) | 4/7 (57.1%) | 67/119 (56.3%) |
| **Experience with suspected or confirmed COVID-19** | | | | | | |
| **Confirmed (by test) cases of COVID-19** (n/N, %)**#**<br>Yes<br><br>Missing | <br><br>228/264 (86.4%)<br>13 | <br><br>50/83 (60.2%)<br>2 | <br><br>5/16 (31.3%)<br><br>1 | <br><br>3/17 (17.6%)<br><br>2 | <br><br>18/58 (31%)<br>1 | <br><br>304/439 (69.2%)+<br><br>19 |
| **Approximate number of confirmed COVID-19 cases**<br>Mean (SD)<br><br>Median (IQR)<br>Total<br>Missing | <br><br>40.2 (59.5)<br><br>14 (5, 52)<br>215<br>62 | <br><br>23 (35.7)<br><br>9 (2.5, 28.5)<br>49<br>36 | <br><br>36.6 (63.5)<br><br>10 (6.5, 80)<br>5<br>12 | <br><br>9 (9.6)<br><br>5 (2, -)<br>3<br>16 | <br><br>6.9 (7.7)<br><br>4 (1, 11)<br>17<br>42 | <br><br>35 (54.7)<br><br>10 (4, 50)<br>289<br>169+ |
| **Services with confirmed COVID-19 cases** (n/N, %)<br>In-patient PC unit<br><br>Hospital PC team<br><br>Home PC team<br><br>Home nursing | <br><br>132/158 (83.5%)<br>117/129 (90.7%)<br>135/151 (89.4%)<br>74/87 (85.1%) | <br><br>26/43 (60.5%)<br>16/26 (61.5%)<br>29/46 (63%)<br><br>7/13 (53.8%) | <br><br>3/8 (37.5%)<br><br>3/9 (33.3%)<br><br>3/9 (33.3%)<br><br>0/1 (0%) | <br><br>1/5 (20%)<br><br>2/9 (22.2%)<br><br>1/7 (14.3%)<br><br>0/3 (0%) | <br><br>10/34 (29.4%)<br>15/36 (41.7%)<br>9/34 (26.5%)<br>3/7 (42.9%) | <br><br>172/249 (69.1%)+<br><br>153/209 (73.2%)<br><br>177/248 (71.4%)+<br><br>84/111 (75.7%) |
| **Suspected cases of COVID-19 (untested but clinical diagnosis/symptoms)** (n/N, %)**#**<br>Missing | 208/260 (80%)<br><br>17 | 46/83 (55.4%)<br><br>2 | 7/17 (41.2%)<br><br><br>- | 6/17 (35.3%)<br><br><br>2 | 27/58 (46.6%)<br><br>1 | 295/436 (67.7%)*<br><br><br>22 |
| **Approximate number of suspected COVID-19 cases**<br>Mean (SD)<br><br>Median (IQR)<br>Total<br>Missing | <br><br>22.9 (42.2)<br><br>10 (4, 21)<br>193<br>84 | <br><br>16.4 (30.4)<br><br>10 (5, 16)<br>47<br>38 | <br><br>20.4 (35.9)<br><br>3 (1, 20)<br>7<br>10 | <br><br>7.7 (7.1)<br><br>7 (1, 12.5)<br>6<br>13 | <br><br>8.3 (10.4)<br><br>5 (2, 10)<br>26<br>33 | <br><br>20 (38)<br><br>10 (4, 20)<br>280+<br>178 |
| **Services with suspected COVID-19 cases** (n/N, %)<br>In-patient PC unit<br><br>Hospital PC team | <br><br>122/156 (78.2%)<br>105/125 (84%) | <br><br>21/43 (48.8%)<br>16/26 (61.5%) | <br><br>2/8 (25%)<br><br>4/9 (44.4%) | <br><br>3/5 (60%)<br><br>3/9 (33.3%) | <br><br>16/34 (47.1%)<br>20/36 (55.6%) | <br><br>165/247 (66.8%)*<br><br>148/205 (72.2%) |

| | | | | | | |
|---|---|---|---|---|---|---|
| Home PC team | 124/150 (82.7%) | 32/46 (69.6%) | 4/10 (40%) | 3/7 (42.9%) | 14/34 (41.2%) | 178/248 (71.8%)* |
| Home nursing | 71/86 (82.6%) | 6/13 (46.2%) | 0/1 (0%) | 0/3 (0%) | 5/7 (71.4%) | 82/110 (74.5%) |
| **Services with confirmed or suspected COVID-19 cases** (n/N, %)# | 248/264 (93.9%) | 60/83 (72.3%) | 9/17 (52.9%) | 7/17 (41.2%) | 33/58 (56.9%) | 358/440 (81.4%)* |
| Missing | 13 | 2 | - | 2 | 1 | 18 |
| **Approximate number of confirmed or suspected COVID-19 cases per service** | | | | | | |
| Median (IQR) | 25.5 (7, 70) | 15 (4.5, 35.5) | 3 (2, 70) | 8 (2, 20) | 6 (2, 11) | 16 (5.5, 56) |
| Total | 234 | 61 | 9 | 7 | 33 | 345+ |
| **Services with confirmed or suspected COVID-19 cases** (n/N, %) | | | | | | |
| In-patient PC unit | 146/158 (92.4%) | 30/43 (69.8%) | 3/8 (37.5%) | 3/5 (60%) | 20/34 (58.8%) | 203/249 (81.5%)* |
| Hospital PC team | 127/129 (98.4%) | 20/26 (76.9%) | 5/9 (55.6%) | 4/9 (44.4%) | 26/36 (72.2%) | 182/209 (87.1%) |
| Home PC team | 143/151 (94.7%) | 35/46 (76.1%) | 5/10 (50%) | 3/7 (42.9%) | 17/34 (50%) | 204/249 (81.9%)* |
| Home nursing | 82/87 (94.3%) | 7/13 (53.8%) | 0/1 (0%) | 0/3 (0%) | 5/7 (71.4%) | 94/111 (84.7%) |
| **Severity of disease in patients with suspected or confirmed COVID-19** (n/N, %) | | | | | | |
| Severely ill or dying due mainly to COVID-19 | 112/248 (45.2%) | 13/60 (21.7%) | 1/9 (11.1%) | 0/7 (0%) | 5/33 (15.2%) | 131/358 (36.6%)+ |
| Pre-existing illnesses/co-morbidities as well as COVID-19 who are severely ill or dying | 192/248 (77.4%) | 34/60 (56.7%) | 3/9 (33.3%) | 5/7 (71.4%) | 15/33 (45.5%) | 250/358 (69.8%)* |
| Patients known to service already who now have COVID-19 | 129/248 (52%) | 21/60 (35%) | 3/9 (33.3%) | 2/7 (28.6%) | 12/33 (36.4%) | 168/358 (46.9%)* |
| **Services who reported that family members/close friends of patients had suspected or confirmed COVID-19** (n/N, %)# | 173/257 (67.3%) | 39/81 (48.1%) | 1/16 (6.3%) | 2/17 (11.8%) | 22/57 (38.6%) | 237/429 (55.2%)+ |
| Missing | 20 | 4 | 1 | 2 | 2 | 29 |
| **Services with staff with suspected or confirmed COVID-19** (n/N, %)# | 238/262 (90.8%) | 55/83 (66.3%) | 4/16 (25%) | 7/17 (41.2%) | 30/58 (51.7%) | 335/437 (76.7%)* |
| Missing | 15 | 2 | 1 | 2 | 1 | 21 |
| **Type of staff with suspected or confirmed COVID-19** (n/N, %) | | | | | | |
| Nurses | 224/238 (94.1%) | 51/55 (92.7%) | 3/4 (75%) | 7/7 (100%) | 22/30 (73.3%) | 308/335 (91.9%)* |
| Physicians | 161/238 (67.6%) | 32/55 (58.2%) | 2/4 (50%) | 3/7 (42.9%) | 15/30 (50%) | 214/335 (63.9%)* |
| Allied health professionals, managed | 92/238 (38.7%) | 11/55 (20%) | 3/4 (75%) | 1/7 (14.3%) | 7/30 (23.3%) | 115/335 (34.3%)* |
| Reception/administrative staff | 74/238 (31.1%) | 9/55 (16.4%) | 0/4 (0%) | 2/7 (28.6%) | 3/30 (10%) | 88/335 (26.3%)+ |
| Managers | 69/238 (29%) | 8/55 (14.5%) | 0/4 (0%) | 1/7 (14.3%) | 0/30 (0%) | 78/335 (23.3%)+ |
| Others | 48/238 (20.2%) | 5/55 (9.1%) | 1/4 (25%) | 0/7 (0%) | 4/30 (13.3%) | 58/335 (17.3%)+ |
| **Services who had volunteers with suspected or confirmed COVID-19** (n/N, %)# | | | | | | |
| Yes | 33/236 (14%) | 3/81 (3.7%) | 1/16 (6.3%) | 0/17 (0%) | 1/56 (1.8%) | 38/407 (9.3%)+ |
| Missing | 41 | 4 | 1 | 2 | 3 | 51 |
| **Service changes in response to COVID-19** | | | | | | |
| **Services who had changed in response to COVID-19** (n/N, %)# | 255/263 (97%) | 74/82 (90.2%) | 14/17 (82.4%) | 15/17 (88.2%) | 53/57 (93%) | 412/437 (94.3%)* |
| Missing | 14 | 3 | - | 2 | 2 | 21 |
| **Services changes in response to COVID-19: level of busyness** (n/N, %) | | | | | | |

| | | | | | | |
|---|---|---|---|---|---|---|
| A lot more busy | 70/255 (27.5%) | 27/74 (36.5%) | 3/14 (21.4%) | 3/15 (20%) | 11/53 (20.8%) | 114/412 (27.7%)+ |
| Slightly more busy | 61/255 (23.9%) | 21/74 (28.4%) | 1/14 (7.1%) | 5/15 (33.3%) | 9/53 (17%) | 97/412 (23.5%)+ |
| About the same | 49/255 (19.2%) | 12/74 (16.2%) | 2/14 (14.3%) | 4/15 (26.7%) | 14/53 (26.4%) | 82/412 (19.9%)* |
| Slightly less busy | 48/255 (18.8%) | 10/74 (13.5%) | 4/14 (28.6%) | 3/15 (20%) | 11/53 (20.8%) | 76/412 (18.4%)+ |
| Much less busy | 27/255 (10.6%) | 4/74 (5.4%) | 4/14 (28.6%) | 0/15 (0%) | 8/53 (15.1%) | 43/412 (10.4%)+ |
| **Services who had lost staff from their service because they have been moved to help the NHS elsewhere** (n/N, %) | 85/253 (33.6%) | 14/73 (19.2%) | 4/13 (30.8%) | 4/15 (26.7%) | 12/53 (22.6%) | 119/408 (29.2%)+ |
| **Services who reported that staff offered to help their service from health services elsewhere** (n/N, %) | 120/253 (47.4%) | 13/73 (17.8%) | 0/13 (0%) | 3/15 (20%) | 9/52 (17.3%) | 145/407 (35.6%)+ |
| **Services who reported changing how their staff work** (n/N, %) | 240/254 (94.5%) | 64/73 (87.7%) | 12/14 (85.7%) | 13/15 (86.7%) | 44/53 (83%) | 374/410 (91.2%)* |
| **Services who reported changing where their staff work (e.g. home working)** (n/N, %) | 217/253 (85.8%) | 43/71 (60.6%) | 7/14 (50%) | 10/15 (66.7%) | 41/53 (77.4%) | 319/407 (78.4%)* |
| **Services who reported changing how their volunteers engage and where** (n/N, %) | 160/223 (71.7%) | 46/71 (64.8%) | 9/13 (69.2%) | 13/15 (86.7%) | 39/51 (76.5%) | 268/374 (71.7%)* |
| **Use of virtual technologies** | | | | | | |
| **Services that reported use of virtual technologies (e.g zoom/teams) with patients and families** (n/N, %) | | | | | | |
| A lot more | 149/252 (59.1%) | 25/71 (35.2%) | 6/13 (46.2%) | 6/15 (40%) | 33/53 (62.3%) | 219/405 (54.1%)+ |
| Slightly more | 71/252 (28.2%) | 24/71 (33.8%) | 2/13 (15.4%) | 8/15 (53.3%) | 14/53 (26.4%) | 120/405 (29.6%)* |
| About the same | 29/252 (11.5%) | 19/71 (26.8%) | 1/13 (7.7%) | 0/15 (0%) | 6/53 (11.3%) | 55/405 (13.6%)+ |
| Slightly less | - | - | - | - | - | - |
| Much less | 3/252 (1.2%) | 3/71 (4.2%) | 4/13 (30.8%) | 1/15 (6.7%) | 0/53 | 11/405 (2.7%)+ |
| **Services that reported use of virtual technologies (e.g zoom/teams) with colleagues** (n/N, %) | | | | | | |
| A lot more | 236/254 (92.9%) | 46/72 (63.9%) | 10/14 (71.4%) | 8/15 (53.3%) | 45/53 (84.9%) | 345/409 (84.4%)+ |
| Slightly more | 14/254 (5.5%) | 14/72 (19.4%) | 3/14 (21.4%) | 5/15 (33.3%) | 8/53 (15.1%) | 45/409 (11%)* |
| About the same | 2/254 (0.8%) | 10/72 (13.9%) | 0/14 | 2/15 (13.3%) | 0/53 | 14/409 (3.4%)+ |
| Slightly less | 1/254 (0.4%) | 2/72 (2.8%) | 0/14 | 0/15 | 0/53 | 3/409 (0.7%)+ |
| Much less | 1/254 (0.4%) | 0/72 | 1/14 (7.1%) | 0/15 | 0/53 | 2/409 (0.5%)+ |
| **Advanced care planning (ACP)** | | | | | | |
| **Services that reported that they are now involved directly with patients/families in ACP** (n/N, %) | | | | | | |
| A lot more | 28/253 (11.1%) | 6/72 (8.3%) | 2/14 (14.3%) | 0/15 (0%) | 3/53 (5.7%) | 39/408 (9.6%)+ |
| Slightly more | 75/253 (29.6%) | 13/72 (18.1%) | 1/14 (7.1%) | 3/15 (20%) | 5/53 (9.4%) | 97/408 (23.8%)+ |
| About the same | 128/253 (50.6%) | 45/72 (62.5%) | 6/14 (42.9%) | 10/15 (66.7%) | 43/53 (81.1%) | 233/408 (57.1%)* |
| Slightly less | 13/253 (5.1%) | 6/72 (8.3%) | 0/14 | 1/15 (6.7%) | 1/53 (1.9%) | 21/408 (5.1%)+ |
| Much less | 9/253 (3.6%) | 2/72 (2.8%) | 5/14 (35.7%) | 1/15 (6.7%) | 1/53 (1.9%) | 18/408 (4.4%)+ |
| **Services that reported that they are now involved advising/supporting others and/or educating about ACP** (n/N, %) | | | | | | |

| | | | | | | |
|---|---|---|---|---|---|---|
| A lot more | 75/253 (29.6%) | 11/70 (15.7%) | 3/14 (21.4%) | 4/15 (26.7%) | 8/53 (15.1%) | 101/406 (24.9%)[+] |
| Slightly more | 84/253 (33.2%) | 19/70 (27.1%) | 1/14 (7.1%) | 4/15 (26.7%) | 11/53 (20.8%) | 120/406 (29.6%)[*] |
| About the same | 75/253 (29.6%) | 35/70 (50%) | 5/14 (35.7%) | 6/15 (40%) | 31/53 (58.5%) | 152/406 (37.4%)[+] |
| Slightly less | 12/253 (4.7%) | 3/70 (4.3%) | 0/14 | 0/15 | 1/53 (1.9%) | 16/406 (3.9%)[+] |
| Much less | 7/253 (2.8%) | 2/70 (2.9%) | 5/14 (35.7%) | 1/15 (6.7%) | 2/53 (3.8%) | 17/406 (4.2%)[+] |
| **Bereavement support** | | | | | | |
| **Bereavement support** (n/N, %) | | | | | | |
| A lot more | 32/193 (16.6%) | 2/53 (3.8%) | 1/13 (7.7%) | 0/12 | 2/41 (4.9%) | 37/313 (11.8%)[+] |
| Slightly more | 49/193 (25.4%) | 14/53 (26.4%) | 1/13 (7.7%) | 5/12 (41.7%) | 9/41 (22%) | 78/313 (24.9%)[+] |
| About the same | 76/193 (39.4%) | 24/53 (45.3%) | 5/13 (38.5%) | 2/12 (16.7%) | 21/41 (51.2%) | 129/313 (41.2%)[*] |
| Slightly less | 23/193 (11.9%) | 9/53 (17%) | 1/13 (7.7%) | 4/12 (33.3%) | 6/41 (14.6%) | 43/313 (13.7%)[+] |
| Much less | 13/193 (6.7%) | 4/53 (7.5%) | 5/13 (38.5%) | 1/12 (8.3%) | 3/41 (7.3%) | 26/313 (8.3%)[+] |
| **Other support** | | | | | | |
| **Services that reported encountering patients or families with COVID-19 who are from black and minority ethnic groups** (n/N, %)# | 93/254 (36.6%) | 16/76 (21.1%) | 3/16 (18.8%) | 5/16 (31.3%) | 15/56 (26.8%) | 132/419 (31.5%)[+] |
| Missing | 23 | 9 | 1 | 3 | 3 | 39 |
| **Changes in specific services** | | | | | | |
| **Services that reported changes in in-patient beds in their service** (n/N, %) | 117/255 (45.9%) | 25/74 (33.8%) | 4/14 (28.6%) | 4/15 (26.7%) | 20/53 (37.7%) | 170/412 (41.3%)[+] |
| **Changes in in-patient beds in your services** | | | | | | |
| Number of beds (n/N, %) | | | | | | |
| Increased | 41/117 (35%) | 5/28 (17.9%) | 1/4 (25%) | 0/4 (0%) | 1/21 (4.8%) | 48/174 (27.6%) |
| Stayed about the same | 34/117 (29.1%) | 10/28 (35.7%) | 2/4 (50%) | 4/4 (100%) | 7/21 (33.3%) | 57/174 (32.8%) |
| Decreased | 42/117 (35.9%) | 13/28 (46.4%) | 1/4 (25%) | 0/4 (0%) | 13/21 (61.9%) | 69/174 (39.7%) |
| **Services that reported changes in how they provide support for patients in acute hospitals** (n/N, %) | 103/255 (40.4%) | 22/74 (29.7%) | 6/14 (42.9%) | 8/15 (53.3%) | 30/53 (56.6%) | 169/412 (41%)[+] |
| **Number of patients needing support in acute hospital** (n/N, %) | | | | | | |
| Increased | 44/101 (43.6%) | 8/25 (32%) | 2/6 (33.3%) | 1/10 (10%) | 9/31 (29%) | 64/173 (37%) |
| Stayed about the same | 28/101 (27.7%) | 4/25 (16%) | 2/6 (33.3%) | 4/10 (40%) | 13/31 (41.9%) | 51/173 (29.5%) |
| Decreased | 29/101 (28.7%) | 13/25 (52%) | 2/6 (33.3%) | 5/10 (50%) | 9/31 (29%) | 58/173 (33.5%) |
| **In acute hospital, face to face contact with patients/their family members is** (n/N, %) | | | | | | |
| A lot more | 9/103 (8.7%) | 2/25 (8%) | 0/6 (0%) | 0/10 (0%) | 0/32 (0%) | 11/176 (6.3%) |
| Slightly more | 6/103 (5.8%) | 2/25 (8%) | 0/6 (0%) | 0/10 (0%) | 2/32 (6.3%) | 10/176 (5.7%) |
| About the same | 14/103 (13.6%) | 2/25 (8%) | 1/6 (16.7%) | 1/10 (10%) | 3/32 (9.4%) | 21/176 (11.9%) |
| Slightly less | 33/103 (32%) | 5/25 (20%) | 1/6 (16.7%) | 3/10 (30%) | 8/32 (25%) | 50/176 (28.4%) |
| Much less | 41/103 (39.8%) | 14/25 (56%) | 4/6 (66.7%) | 6/10 (60%) | 19/32 (59.4%) | 84/176 (47.7%) |
| **In acute hospital, face to face contact with staff is** (n/N, %) | | | | | | |
| A lot more | 29/103 (28.2%) | 2/25 (8%) | 0/6 (0%) | 1/10 (10%) | 0/32 | 32/176 (18.2%) |
| Slightly more | 28/103 (27.2%) | 2/25 (8%) | 0/6 (0%) | 0/10 (0%) | 1/32 (3.1%) | 31/176 (17.6%) |
| About the same | 15/103 (14.6%) | 4/25 (16%) | 0/6 (0%) | 2/10 (20%) | 12/32 (37.5%) | 33/176 (18.8%) |

| | | | | | | |
|---|---|---|---|---|---|---|
| Slightly less | 16/103 (15.5%) | 11/25 (44%) | 3/6 (50%) | 4/10 (40%) | 7/32 (21.9%) | 41/176 (23.3%) |
| Much less | 15/103 (14.6%) | 6/25 (24%) | 3/6 (50%) | 3/10 (30%) | 12/32 (37.5%) | 39/176 (22.2%) |
| **In acute hospital, telephone/remote connection is** (n/N, %) | | | | | | |
| A lot more | 66/104 (63.5%) | 16/25 (64%) | 1/6 (16.7%) | 7/10 (70%) | 17/31 (54.8%) | 107/176 (60.8%) |
| Slightly more | 25/104 (24%) | 8/25 (32%) | 4/6 (66.7%) | 3/10 (30%) | 11/31 (35.5%) | 51/176 (29%) |
| About the same | 11/104 (10.6%) | 1/25 (4%) | 1/6 (16.7%) | 0/10 (0%) | 3/31 (9.7%) | 16/176 (9.1%) |
| Slightly less | 1/104 (1%) | 0/25 (0%) | 0/6 (0%) | 0/10 (0%) | 0/31 | 1/176 (0.6%) |
| Much less | 1/104 (1%) | 0/25 (0%) | 0/6 (0%) | 0/10 (0%) | 0/31 | 1/176 (0.6%) |
| **Services that reported changes in how they provide support for patients in their own homes** (n/N, %) | 152/255 (59.6%) | 32/74 (43.2%) | 8/14 (57.1%) | 6/15 (40%) | 30/53 (56.6%) | 228/412 (55.3%)+ |
| **Number of patients needing support in their own homes** (n/N, %) | | | | | | |
| Increased | 60/152 (39.5%) | 15/32 (46.9%) | 1/8 (12.5%) | 4/6 (66.7%) | 13/31 (41.9%) | 93/229 (40.6%) |
| Stayed about the same | 72/152 (47.4%) | 11/32 (34.4%) | 4/8 (50%) | 1/6 (16.7%) | 15/31 (48.4%) | 103/229 (45%) |
| Decreased | 20/152 (13.2%) | 6/32 (18.8%) | 3/8 (37.5%) | 1/6 (16.7%) | 3/31 (9.7%) | 33/229 (14.4%) |
| **Face to face contact with patients/their family members in their own homes is** (n/N, %) | | | | | | |
| A lot more | 4/153 (2.6%) | 1/31 (3.2%) | 1/8 (12.5%) | 0/6 (0%) | 3/31 (9.7%) | 9/229 (3.9%) |
| Slightly more | 2/153 (1.3%) | 4/31 (12.9%) | 0/8 (0%) | 0/6 (0%) | 2/31 (6.5%) | 8/229 (3.5%) |
| About the same | 8/153 (5.2%) | 5/31 (16.1%) | 1/8 (12.5%) | 1/6 (16.7%) | 1/31 (3.2%) | 16/229 (7%) |
| Slightly less | 38/153 (24.8%) | 6/31 (19.4%) | 2/8 (25%) | 1/6 (16.7%) | 9/31 (29%) | 56/229 (24.5%) |
| Much less | 101/153 (66%) | 15/31 (48.4%) | 4/8 (50%) | 4/6 (66.7%) | 16/31 (51.6%) | 140/229 (61.1%) |
| **Face to face contact with staff in patients' homes is** (n/N, %) | | | | | | |
| A lot more | 2/150 (1.3%) | 0/31 (0%) | 1/8 (12.5%) | 0/6 (0%) | 1/29 (3.4%) | 4/224 (1.8%) |
| Slightly more | 5/150 (3.3%) | 2/31 (6.5%) | 0/8 (0%) | 0/6 (0%) | 1/29 (3.4%) | 8/224 (3.6%) |
| About the same | 24/150 (16%) | 5/31 (16.1%) | 3/8 (37.5%) | 1/6 (16.7%) | 5/29 (17.2%) | 38/224 (17%) |
| Slightly less | 48/150 (32%) | 9/31 (29%) | 2/8 (25%) | 2/6 (33.3%) | 10/29 (34.5%) | 71/224 (31.7%) |
| Much less | 71/150 (47.3%) | 15/31 (48.4%) | 2/8 (25%) | 3/6 (50%) | 12/29 (41.4%) | 103/224 (46%) |
| **For patients in their own homes, telephone/remote connection is** (n/N, %) | | | | | | |
| A lot more | 125/151 (82.8%) | 21/32 (65.6%) | 2/8 (25%) | 5/6 (83.3%) | 22/31 (71%) | 175/228 (76.8%) |
| Slightly more | 20/151 (13.2%) | 9/32 (28.1%) | 4/8 (50%) | 0/6 (0%) | 6/31 (19.4%) | 39/228 (17.1%) |
| About the same | 6/151 (4%) | 2/32 (6.3%) | 1/8 (12.5%) | 1/6 (16.7%) | 2/31 (6.5%) | 12/228 (5.3%) |
| Much less | 0/151 (0%) | 0/32 (0%) | 1/8 (12.5%) | 0/6 (0%) | 1/31 (3.2%) | 2/228 (0.9%) |
| **Services that reported changes in how medicines are given in the community (e.g. who sets up syringe drivers/families administering medicines** (n/N, %) | 56/149 (37.6%) | 7/31 (22.6%) | 1/8 (12.5%) | 2/6 (33.3%) | 9/28 (32.1%) | 75/222 (33.8%) |
| **Services that reported changes in how they provide support for patients in care homes (including nursing homes)** (n/N, %) | 96/255 (37.6%) | 23/74 (31.1%) | 2/14 (14.3%) | 4/15 (26.7%) | 28/53 (52.8%) | 153/412 (37.1%)+ |

| | | | | | | |
|---|---|---|---|---|---|---|
| **Number of patients needing support in care homes (including nursing homes)** (n/N, %) | | | | | | |
| Increased | 40/93 (43%) | 7/23 (30.4%) | 0/2 (0%) | 2/3 (66.7%) | 5/25 (20%) | 54/146 (37%) |
| Stayed about the same | 45/93 (48.4%) | 7/23 (30.4%) | 1/2 (50%) | 1/3 (33.3%) | 14/25 (56%) | 68/146 (46.6%) |
| Decreased | 8/93 (8.6%) | 9/23 (39.1%) | 1/2 (50%) | 0/3 (0%) | 6/25 (24%) | 24/146 (16.4%) |
| **Face to face contact with patients/their family members in care homes (including nursing homes) is** (n/N, %) | | | | | | |
| A lot more | 6/94 (6.4%) | 3/21 (14.3%) | 0/2 (0%) | 0/3 (0%) | 0/25 (0%) | 9/145 (6.2%) |
| Slightly more | 6/94 (6.4%) | 0/21 (0%) | 0/2 (0%) | 0/3 (0%) | 1/25 (4%) | 7/145 (4.8%) |
| About the same | 12/94 (12.8%) | 0/21 (0%) | 1/2 (50%) | 0/3 (0%) | 0/25 (0%) | 13/145 (9%) |
| Slightly less | 18/94 (19.1%) | 5/21 (23.8%) | 0/2 (0%) | 2/3 (66.7%) | 4/25 (16%) | 29/145 (20%) |
| Much less | 52/94 (55.3%) | 13/21 (61.9%) | 1/2 (50%) | 1/3 (33.3%) | 20/25 (80%) | 87/145 (60%) |
| **Face to face contact with staff in care homes (including nursing homes) is** (n/N, %) | | | | | | |
| A lot more | 8/93 (8.6%) | 2/21 (9.5%) | 0/2 (0%) | 2/3 (66.7%) | 0/25 | 12/144 (8.3%) |
| Slightly more | 7/93 (7.5%) | 2/21 (9.5%) | 0/2 (0%) | 0/3 (0%) | 1/25 (4%) | 10/144 (6.9%) |
| About the same | 7/93 (7.5%) | 3/21 (14.3%) | 1/2 (50%) | 1/3 (33.3%) | 2/25 (8%) | 14/144 (9.7%) |
| Slightly less | 15/93 (16.1%) | 2/21 (9.5%) | 1/2 (50%) | 0/3 (0%) | 4/25 (16%) | 22/144 (15.3%) |
| Much less | 56/93 (60.2%) | 12/21 (57.1%) | 0/2 (0%) | 0/3 (0%) | 18/25 (72%) | 86/144 (59.7%) |
| **For patients in care homes (including nursing homes), telephone/remote connection is** (n/N, %) | | | | | | |
| A lot more | 57/93 (61.3%) | 14/23 (60.9%) | 0/2 (0%) | 2/3 (66.7%) | 10/25 (40%) | 83/146 (56.8%) |
| Slightly more | 20/93 (21.5%) | 6/23 (26.1%) | 1/2 (50%) | 1/3 (33.3%) | 7/25 (28%) | 35/146 (24%) |
| About the same | 11/93 (11.8%) | 2/23 (8.7%) | 0/2 (0%) | 0/3 (0%) | 7/25 (28%) | 20/146 (13.7%) |
| Slightly less | 4/93 (4.3%) | 1/23 (4.3%) | 1/2 (50%) | 0/3 (0%) | 1/25 (4%) | 7/146 (4.8%) |
| Much less | 1/93 (1.1%) | 0/23 (0%) | 0/2 (0%) | 0/3 (0%) | 0/25 (0%) | 1/146 (0.7%) |
| **Changes to how you are supporting families/those important to patients** | | | | | | |
| **Support for families/ those important to patients compared to before is** (n/N, %) | | | | | | |
| A lot more | 43/247 (17.4%) | 5/66 (7.6%) | 1/13 (7.7%) | 2/15 (13.3%) | 12/47 (25.5%) | 63/388 (16.2%) |
| Slightly more | 49/247 (19.8%) | 16/66 (24.2%) | 3/13 (23.1%) | 3/15 (20%) | 5/47 (10.6%) | 76/388 (19.6%) |
| About the same | 82/247 (33.2%) | 24/66 (36.4%) | 3/13 (23.1%) | 5/15 (33.3%) | 18/47 (38.3%) | 132/388 (34%) |
| Slightly less | 57/247 (23.1%) | 14/66 (21.2%) | 2/13 (15.4%) | 3/15 (20%) | 6/47 (12.8%) | 82/388 (21.1%) |
| Much less | 16/247 (6.5%) | 7/66 (10.6%) | 4/13 (30.8%) | 2/15 (13.3%) | 6/47 (12.8%) | 35/388 (9%) |
| **Services that reported that they had changed how they contact and work with families/those important to patients** (n/N, %) | 217/243 (89.3%) | 44/66 (66.7%) | 10/13 (76.9%) | 11/14 (78.6%) | 37/48 (77.1%) | 319/384 (83.1%) |
| **Changes to how you are deploying volunteers** | | | | | | |
| **Services that reported changes to how they deploy volunteers** (n/N, %) | 155/194 (79.9%) | 29/51 (56.9%) | 6/10 (60%) | 13/15 (86.7%) | 33/43 (76.7%) | 236/313 (75.4%) |
| **Deployment of volunteers compared to before is** (n/N, %) | | | | | | |
| A lot more | 5/157 (3.2%) | 3/32 (9.4%) | 0/7 (0%) | 0/14 | 2/34 (5.9%) | 10/244 (4.1%) |
| Slightly more | 4/157 (2.5%) | 2/32 (6.3%) | 0/7 (0%) | 2/14 (14.3%) | 0/34 (0%) | 8/244 (3.3%) |
| About the same | 7/157 (4.5%) | 1/32 (3.1%) | 0/7 (0%) | 0/14 | 1/34 (2.9%) | 9/244 (3.7%) |
| Slightly less | 15/157 (9.6%) | 2/32 (6.3%) | 3/7 (42.9%) | 1/14 (7.1%) | 3/28 (8.8%) | 24/244 (9.8%) |
| Much less | 126/157 (80.3%) | 24/32 (75%) | 4/7 (57.1%) | 11/14 (78.6%) | 28/34 (82.4%) | 193/244 (79.1%) |
| **Shortages** | | | | | | |

| | | | | | | |
|---|---|---|---|---|---|---|
| **Personal Protective Equipment (PPE) shortages** (n/N, %)#<br>Missing or not applicable[£] | 129/258 (50%)<br>148 | 38/75 (50.7%)<br>47 | 9/15 (60%)<br>8 | 6/17 (35.3%)<br>13 | 19/54 (35.2%)<br>40 | 201/419 (48%)<br>256 |
| **PPE shortages in the last 7 days[&]** (n/N, %)#<br>Missing or not applicable[£] | 50/127 (39.4%)<br>227 | 7/38 (18.4%)<br>78 | 7/9 (77.8%)<br>10 | 5/6 (83.3%)<br>14 | 6/19 (31.6%)<br>53 | 75/199 (37.7%)<br>382 |
| **Key medicines shortages** (n/N, %)#<br>Missing or not applicable[£] | 63/255 (24.7%)<br>214 | 19/73 (26%)<br>66 | 4/15 (26.7%)<br>13 | 4/17 (23.5%)<br>15 | 11/54 (20.4%)<br>48 | 101/414 (24.4%)<br>356 |
| **Key medicines shortages in the last 7 days[&]** (n/N, %)#<br>Missing or not applicable[£] | 28/63 (44.4%)<br>249 | 3/18 (16.7%)<br>82 | 4/4 (100%)<br>13 | 4/4 (100%)<br>15 | 6/11 (54.5%)<br>53 | 45/100 (45%)<br>412 |
| **Equipment shortages (e.g. syringe drivers)** (n/N, %)#<br>Missing or not applicable[£] | 45/256 (17.6%)<br>232 | 3/73 (4.1%)<br>82 | 4/14 (28.6%)<br>13 | 2/17 (11.8%)<br>17 | 2/54 (3.7%)<br>57 | 56/414 (13.5%)<br>401 |
| **Equipment shortages in the last 7 days[&]** (n/N, %)#<br>Missing or not applicable[£] | 19/42 (45.2%)<br>258 | 1/3 (33.3%)<br>84 | 3/3 (100%)<br>14 | 2/2 (100%)<br>17 | 2/2 (100%)<br>57 | 27/52 (51.9%)<br>430 |
| **Staff shortages** (n/N, %)#<br>Missing or not applicable[£] | 117/255 (45.9%)<br>160 | 26/75 (34.7%)<br>59 | 4/14 (28.6%)<br>13 | 6/16 (37.5%)<br>13 | 13/54 (24.1%)<br>46 | 166/414 (40.1%)<br>291 |
| **Staff shortages in the last 7 days[&]** (n/N, %)#<br>Missing or not applicable[£] | 61/114 (53.5%)<br>216 | 9/26 (34.6%)<br>76 | 3/4 (75%)<br>14 | 5/5 (100%)<br>14 | 5/13 (38.5%)<br>54 | 83/162 (51.2%)<br>374 |

59 of the 458 responses completed data but did not hit the final submit button, so lack a date and time stamp. Many were contacted and had no further information. Only 4/458 responders chose not to provide an email address on registration.

Note: UK = United Kingdom, Rest of Europe excludes UK, LIC = Low Income Countries, LMIC = Lower Middle Income Countries, UMIC = Upper Middle Income Countries, HIC = High Income countries, IQR = Interquartile range, NHS = National Health Service, PC = Palliative Care

### n of value and valid N denominator are provided. Percentages are of valid values, unless otherwise stated. Number of missing responses for each category are provided below. * Included data from the one missing country in the numerator and denominator. [+] Includes data from the one missing country in the denominator.

[&]Response for "yes" and "sometimes" both coded as "yes"

[£]Not applicable includes those services who did not have shortages

##### Table S 2 Models of care in overall, charity/non-profit and public palliative care settings and numbers of services at only one, only two, only three or all of the settings

| Settings and Setting Combinations only at the stated settings | Numbers of services according to management type at specified settings* | | | |
|---|---|---|---|---|
| | **Overall (n=458)** | **Charity/ Non-Profit (n=192)** | **Public (n=204)** | **Other/Private/Unspecified (n=62)** |
| **Inpatient Palliative Care Unit** | 59 | 31 | 19 | 9 |
| Inpatient Palliative Care Unit & Hospital Palliative Care Team | 23 | 6 | 12 | 5 |
| Inpatient Palliative Care Unit & Home Palliative Care Team | 40 | 31 | 5 | 4 |
| Inpatient Palliative Care Unit & Home Nursing | 14 | 13 | 0 | 1 |
| Inpatient Palliative Care Unit & Home Palliative Care Team & Home Nursing | 47 | 42 | 1 | 4 |
| Inpatient Palliative Care Unit & Hospital Palliative Care Team & Home Nursing | 1 | 1 | 0 | 0 |
| Inpatient Palliative Care Unit & Hospital Palliative Care Team & Home Palliative Care Team | 53 | 12 | 27 | 14 |
| Inpatient Palliative Care Unit & Hospital Palliative Care Team & Home Palliative Care Team & Home Nursing | 24 | 18 | 3 | 3 |
| **Hospital Palliative Care Team** | 85 | 2 | 77 | 6 |
| Hospital Palliative Care Team & Home Palliative Care Team | 26 | 0 | 21 | 5 |

| | | | | |
|---|---|---|---|---|
| Hospital Palliative Care Team & Home Nursing | 0 | 0 | 0 | 0 |
| Hospital Palliative Care Team & Home Palliative Care Team & Home Nursing | 5 | 1 | 3 | 1 |
| **Home Palliative Care Team** | 48 | 16 | 25 | 7 |
| Home Palliative Care Team & Home Nursing | 18 | 11 | 5 | 2 |
| **Home Nursing** | 10 | 6 | 4 | 0 |

*Data is missing for 2 Non-Profit, 2 Public and 1 Other/Private/Unspecified service

#### Table S 3 Overall impact of COVID-19 on palliative care services, main themes and example quotes

**IMPACT OF COVID-19 ON PALLIATIVE CARE SERVICES**

**EMOTIONAL IMPACT**

**Fatigue**

*'Emotional fatigue because of a different type of dying & volume, as well as a lack of other type of work'* UK, Other (part of an NHS acute trust)

'*fatigue - at times - with rapid changes and maintaining a flexible response to change'* UK, Charitable/Non-Profit

*'Volunteer death, staff family members' severe illness and also one death, and a local GP's death have hit hard. Fatigue has been a feature …tearfulness and also staff not wanting to be away/wanting to give.'* UK, Charitable/Non-Profit

*'Very intense way of working - no separation of work and home environment- radical changes to practice and assurance of retention of quality and safety for staff and patients has been very demanding…'* Rest of Europe, Charitable/Non-Profit

**Anxiety and Worry**

*'…staff have been anxious about their own health and the health of their families…have suffered bereavements and some have been hospitalised themselves, this undoubtedly has an impact on team morale'* UK, Charitable/Non-profit

*'…some normally resilient and balanced staff have been affected psychologically by health anxieties and not able to function or asking to be redeployed into non patient facing roles.'* UK, Charitable/Non-Profit

*'…just the feeling of I can be dangerous for patients in palliative care, because most time I am working in acute hospital...'* Rest of Europe, Public

**Stress & Distress**

*'…great moral distress in supporting families who are stressed or bereaved that they cannot or could not be with their loved one through death or visit while the client could still communicate.'* Rest of the World (HIC), Public

*'Emotional burden on staff… ethical dilemmas… tested resilience, in some cases eroded'* UK, Charitable/Non-Profit

*'Working with the on-going uncertainty and adjusting to new ways of working has put an emotional strain and pressure on staff, but it has also provided a strength of purpose and commitment.'* UK, Charitable/Non-Profit

*'Staff are more concerned about how to provide care safely and upset that we are not able to offer the same care we would normally'* UK, Charitable/Non-Profit

*'Increased stress has been evident requiring additional support from clinical psychology'* UK, Charitable/Non-Profit

*'Chronic strain re emotional/social restrictions, constant change and uncertainty'* UK, Charitable/Non-Profit

**Guilt**

| |
|---|
| *'For all that I have been less busy than my acute colleagues - and I feel guilty about that …a kind of reverse survivor's guilt…'* UK, Charitable/Non-Profit<br><br>*'Staff members becoming ill generates lots of fear and anxiety. Staff feeling guilty if they think they have passed on Covid to patients.'* UK, Public |
| **FRUSTRATION**<br><br>**Changing policy and practice**<br><br>*'Increased anxiety, and needing to ensure clear communication of changes in policy or practice'* UK, Charitable/Non-Profit<br><br>*'…there has been such difficulty coping with the information overload, sifting through what is essential and what is not. The bed capacity tracker has been a very laborious, time consuming and stressful exercise for all involved but at the same time a necessary evil in order to ensure funding.'* UK, Charitable/Non-Profit<br><br>*'…with the ever changing guidelines, just become familiar and they change again. Changing the service provision and supporting staff through this …everyone is struggling, so it is really difficult to see who to go to for support'* UK, Charitable/Non-Profit<br><br>*'…there was a lot of irritation and insecurity, the regional government changed directives nearly every day, because nobody knew what was best to do. We had a lot of work to implement the frequent changing guideline into our hospital and on the same time to teach the staff appropriately.'* Rest of Europe, Public<br><br>**Lack of Sympathy**<br>*'…lack of understanding of acute colleagues of the difficulties facing palliative providers in the community.'* Rest of the World (HIC), Public<br><br>*'…staff on furlough and the animosity this has caused with staff who are still working…'* UK, Charitable/Non-Profit |
| **FINANCIAL**<br><br>*'Financial distress of hospice resulting in staff taking voluntary pay cuts of up to 30%...'* Rest of the World (UMIC), Charitable/Non-Profit<br><br>*'…fear for oneself and ones patients uncertainty of having a job after this no funding how to sustain the organization as it is a NGO…'* Rest of the World (UMIC), Charitable/Non-Profit |

##### Table S 4 Univariable analysis of potential predictors of PPE shortages

| | | Odds Ratio | 95% CI | | p-value | Number of responses in model |
|---|---|---|---|---|---|---|
| **Unit management type** | Public | Ref | | | | 415 |
| | Charitable | 5.91 | 3.78 | 9.24 | <0.001 | |
| | Other | 1.39 | 0.69 | 2.82 | 0.36 | |
| **Type of patients** | Adults only | Ref | | | | 413 |
| | Mixed | 1.35 | 0.74 | 2.48 | 0.33 | |
| | Children only | 1.15 | 0.52 | 2.54 | 0.74 | |
| **Busyness** | Same | Ref | | | | 419 |
| | A lot more busy | 0.91 | 0.52 | 1.59 | 0.73 | |
| | Slightly more busy | 0.57 | 0.32 | 1.02 | 0.06 | |
| | Slightly less busy | 0.75 | 0.41 | 1.37 | 0.34 | |
| | Much less busy | 0.32 | 0.15 | 0.69 | 0.003 | |
| **Country** | UK | Ref | | | | 419 |
| | Rest of Europe | 1.03 | 0.61 | 1.72 | 0.92 | |
| | Rest of the World | 0.65 | 0.40 | 1.07 | 0.10 | |
| **Any suspected or confirmed COVID patients** | No | Ref | | | | 416 |
| | Yes | 1.20 | 0.73 | 1.97 | 0.47 | |
| **Inpatient palliative care unit** | No | Ref | | | | 419 |
| | Yes | 3.95 | 2.61 | 5.96 | <0.001 | |
| **Hospital palliative care team** | No | Ref | | | | 419 |
| | Yes | 0.29 | 0.20 | 0.44 | <0.001 | |
| **Home palliative care team** | No | Ref | | | | 419 |
| | Yes | 1.58 | 1.07 | 2.33 | 0.02 | |
| **Home nursing** | No | Ref | | | | 419 |
| | Yes | 3.53 | 2.21 | 5.66 | <0.001 | |

Note Ref = reference category

#### Table S 5 Univariable analysis of potential predictors of medicines shortages

| | | Odds Ratio | 95% CI | | p-value | Number of responses in model |
|---|---|---|---|---|---|---|
| **Unit management type** | Public | Ref | | | | 410 |
| | Charitable | 0.73 | 0.45 | 1.18 | 0.20 | |
| | Other | 1.03 | 0.48 | 2.22 | 0.94 | |
| **Type of patients** | Adults only | Ref | | | | 408 |
| | Mixed | 0.68 | 0.31 | 1.45 | 0.32 | |
| | Children only | 0.53 | 0.18 | 1.59 | 0.26 | |
| **Busyness** | Same | Ref | | | | 414 |
| | A lot more busy | 1.24 | 0.66 | 2.35 | 0.50 | |
| | Slightly more busy | 1.12 | 0.58 | 2.17 | 0.74 | |
| | Slightly less busy | 0.81 | 0.39 | 1.68 | 0.57 | |
| | Much less busy | 0.57 | 0.22 | 1.46 | 0.24 | |
| **Country** | UK | Ref | | | | 414 |
| | Rest of Europe | 1.07 | 0.59 | 1.94 | 0.82 | |
| | Rest of the World | 0.86 | 0.48 | 1.55 | 0.62 | |
| **Any suspected or confirmed COVID patients** | No | Ref | | | | 411 |
| | Yes | 1.36 | 0.73 | 2.52 | 0.33 | |
| **Inpatient palliative care unit** | No | Ref | | | | 414 |
| | Yes | 0.66 | 0.42 | 1.04 | 0.07 | |
| **Hospital palliative care team** | No | Ref | | | | 414 |
| | Yes | 1.23 | 0.78 | 1.93 | 0.37 | |
| **Home palliative care team** | No | Ref | | | | 414 |
| | Yes | 1.34 | 0.85 | 2.11 | 0.21 | |
| **Home nursing** | No | Ref | | | | 414 |
| | Yes | 0.70 | 0.41 | 1.21 | 0.20 | |

Note: Ref = reference category

##### Table S 6 Univariable analysis of potential predictors of other equipment shortages

| | | Odds Ratio | 95% CI | | p-value | Number of responses in model |
|---|---|---|---|---|---|---|
| **Unit management type** | Public | Ref | | | | 410 |
| | Charitable | 0.56 | 0.31 | 1.04 | 0.07 | |
| | Other | 0.51 | 0.17 | 1.54 | 0.24 | |
| **Type of patients** | Adults only | Ref | | | | 408 |
| | Mixed | 0.57 | 0.20 | 1.67 | 0.31 | |
| | Children only | 0.50 | 0.11 | 2.18 | 0.36 | |
| **Busyness** | Same | Ref | | | | 414 |
| | A lot more busy | 10.27 | 3.01 | 35.07 | <0.001 | |
| | Slightly more busy | 5.57 | 1.55 | 19.96 | 0.008 | |
| | Slightly less busy | 3.41 | 0.87 | 13.30 | 0.08 | |
| | Much less busy | 1.40 | 0.22 | 8.68 | 0.72 | |
| **Country** | UK | Ref | | | | 414 |
| | Rest of Europe | 0.20 | 0.06 | 0.67 | 0.009 | |
| | Rest of the World | 0.49 | 0.22 | 1.08 | 0.08 | |
| **Any suspected or confirmed COVID patients** | No | Ref | | | | 411 |
| | Yes | 1.88 | 0.77 | 4.58 | 0.16 | |
| **Inpatient palliative care unit** | No | Ref | | | | 414 |
| | Yes | 0.38 | 0.21 | 0.69 | 0.001 | |
| **Hospital palliative care team** | No | Ref | | | | 414 |
| | Yes | 1.20 | 0.68 | 2.11 | 0.52 | |
| **Home palliative care team** | No | Ref | | | | 414 |
| | Yes | 0.88 | 0.50 | 1.55 | 0.66 | |
| **Home nursing** | No | Ref | | | | 414 |
| | Yes | 0.77 | 0.39 | 1.51 | 0.44 | |

Note: Ref = reference category

##### Table S 7 Univariable analysis of potential predictors of staff shortages

| | | Odds Ratio | 95% CI | | p-value | Number of responses in model |
|---|---|---|---|---|---|---|
| **Unit management type** | Public | Ref | | | | 410 |
| | Charitable | 1.33 | 0.88 | 2.01 | 0.19 | |
| | Other | 0.96 | 0.47 | 1.97 | 0.91 | |
| **Type of patients** | Adults only | Ref | | | | 408 |
| | Mixed | 0.46 | 0.23 | 0.92 | 0.03 | |
| | Children only | 0.24 | 0.08 | 0.70 | 0.009 | |
| **Busyness** | Same | Ref | | | | 414 |
| | A lot more busy | 3.18 | 1.76 | 5.74 | <0.001 | |
| | Slightly more busy | 1.60 | 0.86 | 2.96 | 0.14 | |
| | Slightly less busy | 1.52 | 0.79 | 2.91 | 0.21 | |
| | Much less busy | 0.92 | 0.41 | 2.06 | 0.85 | |
| **Country** | UK | Ref | | | | 414 |
| | Rest of Europe | 0.63 | 0.37 | 1.07 | 0.09 | |
| | Rest of the World | 0.44 | 0.26 | 0.76 | 0.003 | |
| **Any suspected or confirmed COVID patients** | No | Ref | | | | 411 |
| | Yes | 2.54 | 1.41 | 4.54 | 0.02 | |
| **Inpatient palliative care unit** | No | Ref | | | | 414 |
| | Yes | 1.34 | 0.90 | 2.00 | 0.15 | |
| **Hospital palliative care team** | No | Ref | | | | 414 |
| | Yes | 0.96 | 0.65 | 1.42 | 0.84 | |
| **Home palliative care team** | No | Ref | | | | 414 |
| | Yes | 1.08 | 0.73 | 1.61 | 0.69 | |
| **Home nursing** | No | Ref | | | | 414 |
| | Yes | 1.30 | 0.83 | 2.03 | 0.24 | |

Note: Ref = Reference category

**Appendix II**

CovPall Questionnaire

### COVPALL COLLABORATION SURVEY

Improving palliative care for people with COVID-19 by sharing learning

Thank you for agreeing to complete this survey. We are trying to find out about how palliative care and hospice services are changing as a result of the COVID-19 pandemic. This is important as the disease is new and hospices/palliative care services are changing how they work and there is an opportunity to learn from each other.

We realise you are very busy right now, and so we have tried to balance collecting the information that patients, policy makers and services think is most helpful, with keeping the questionnaire as short as we can.

The questionnaire has 7 sections, and should take no longer than 30 minutes to complete, although it may depend on how much additional/open comments you wish to share. We will consider everything that you say. Your reply will help us. Although the grouped results will be shared, we will not name your unit unless you ask us to.

You can pause the questionnaire by clicking the "Save & Return Later" button at the bottom of each page. You will be given a code to enable you to continue later. (There should be a "returning" option at the top right of this page.) If you wish to correct errors after you have clicked "Submit" please with CovPall in the subject line.

If you would like help in completing the survey, or would prefer someone to read the questions to you while you give the answers over the telephone / zoom call, please send your contact details to with CovPall in the subject line.

If you have any concerns about this questionnaire or this study please . CovPall is led by Professor Irene Higginson of the Cicely Saunders Institute, with a multiprofessional team of partners from different organisations and backgrounds. Patient representatives have contributed to this questionnaire and our plans. For more information see https://www.kcl.ac.uk/cicelysaunders (specific page later).

This study has been granted ethical approval by the PNM Research Ethics Subcommittee of King's College London, code LRS-19/20-18541.

Please tick all responses that apply.

#### 1. ABOUT YOU AND YOUR SERVICE

1.1 Contact email of the person completing the survey ________________________________
(We need this information so we can help you get back into the survey, or help you to complete it, if you have difficulty)

1.2 Name of the person completing the survey ________________________________

1.3 Date ________________________________
(DD-MM-YYYY)

1.4 Country

◯ England
◯ Scotland
◯ Wales
◯ N Ireland
◯ Australia
◯ Belgium
◯ Canada
◯ Germany
◯ Ireland
◯ Italy
◯ Poland
◯ New Zealand
◯ Other (a box will open)
(A regions option may appear)

1.4a Country (please specify) ________________________________

1.4b English Regions

◯ North East
◯ North West
◯ Yorkshire and The Humber
◯ East Midlands
◯ West Midlands
◯ East
◯ London
◯ South East
◯ South West

1.4c Welsh region

◯ N Wales
◯ W Wales
◯ SE Wales

1.4d Scottish region

◯ Fife, Lothian, Borders, Dumfries & Galloway
◯ Greater Glasgow & Clyde, Ayrshire & Arran, Lanarkshire, Forth Valley
◯ Tayside, Grampian, Western Isles, Highland, Orkney and Shetland

1.4e Region ________________________________

1.5 Your role

◯ Medical director / lead medical clinician
◯ Nurse director / lead nurse clinician
◯ Other (a box will open below)

1.5a Please specify your role ________________________________

#### 2. ABOUT THE SERVICES YOU USUALLY OFFERED BEFORE THE COVID-19 PANDEMIC.
**Please tick all answers that apply unless indicted**

2.1 Types of patients cared for

☐ Adults ☐ Children

2.2 In what settings did you provide palliative care services

(additional questions will open for each choice)

☐ In-patient hospice / palliative care unit
☐ Hospital palliative care advisory team
☐ Specialist palliative home care service (supporting / consulting about care for patients at home and/or in the community)
☐ Providing hands on nursing care at home / in the community (e.g. hospice@home, pall@home)
(Tick all that apply)

#### Questions for in-patient hospice / palliative care unit

2.2a Number of beds ______________ (Must be a number)

2.2b Approximate number of new patients seen annually ______________ (Must be a number)

2.2c Normal hours of admitting patients ______________ (E.g. 9:30-17:00)

#### Questions for hospital palliative care advisory team

2.2d Approximate number of new patients seen annually ______________ (Must be a number)

2.2e Did you support
- ☐ Acute hospitals
- ☐ Community hospitals

2.2f Normal hours of accepting new referrals ______________ (E.g. 9:30-17:00)

2.2g Did you offer 24/7 support for your patients ☐ Yes

#### Questions for specialist palliative home care service

2.2h Approximate number of new patients seen annually ______________ (Must be a number)

2.2i Did you support patients in care homes ○ Yes ○ No

2.2j Normal hours of accepting new referrals ______________ (E.g. 9:30-17:00)

2.2k Did you offer 24/7 support for your patients ☐ Yes

#### Questions for providing hands on nursing care at home / in the community

2.2l Approximate number of new patients seen annually ______________ (Must be a number)

2.2m Normal hours of accepting new referrals ______________ (E.g. 9:30-17:00)

2.2n Did you offer 24/7 support for your patients ☐ Yes

#### Additional information about services offered before the COVID-19 pandemic

2.3 Normal hours of accepting new referrals ________________________________

2.4 Did you offer bereavement services ◯ Yes ◯ No

(if yes - additional questions will open)

2.4a Bereavement services / support usually provided and to whom ________________________________

2.4b Did you offer bereavement services only to families / friends of patients who had been cared for by your service ◯ Yes ◯ No

2.4c Whom did you offer support to ________________________________

2.4d Did you use a risk assessment tool to help you decide how to target bereavement services ◯ Yes ◯ No

2.5 Other services provided, e.g. Day care, rehabilitation, lymphoedema, outpatient clinics - please detail ________________________________

2.6 If you had volunteer roles available within your service, what were they

Tick all that apply

- ☐ Direct patient / family facing support (e.g. befriending, home visits, in-patient unit care, family support groups / visiting etc.)
- ☐ Indirect patient / family facing support (e.g. reception functions, refreshments, driving / transport etc.)
- ☐ Back office functions (e.g. finance support, maintenance, gardening etc.)
- ☐ Fundraising functions (e.g. shop volunteers, lottery etc.)
- ☐ Others (a box will open below)

2.6a Please specify the other volunteer roles ________________________________

2.7 Did you use remote consultations to help support patients in your care or for education before the COVID-19 Pandemic

Tick all that apply (an example box will appear)

- ☐ Telephone support for education
- ☐ Telephone support for clinical care
- ☐ Telehealth / video support / e-learning for education
- ☐ Telehealth / video support / e-learning for clinical care

2.7a Please give a brief example of the use of remote consultations ________________________________

2.8 Is your service managed as a unit that is

- ◯ Charitable / non-profit
- ◯ Public
- ◯ Private
- ◯ Other (a box will open below)

2.8a Please explain your unit type ________________________________

2.8b what percentage of your funding was usually from the NHS ________________________________
(Must be a number (0-100))

2.9 How well would you say your service was integrated with other NHS primary or secondary care services in your area (e.g. with hospitals, primary care etc.)

Please rank the level of integration: from 0 (no integration at all, no connections) to 10 (very well integrated / close working and planning)

0 5 10

(Place a mark on the scale above)

2.10 Any comments on integration ______________________

2.11 Is there anything else you want to tell us about how your service operates that you think is important for us to know ______________________

#### 3. EXPERIENCE WITH SUSPECTED OR CONFIRMED CASES OF COVID-19. Please tick all that apply

**Please tell us about people you have encountered with suspected or confirmed COVID-19**

3.1 Have you had any patients with confirmed (by test) cases of COVID-19

(additional questions will open if you tick yes)

○ Yes ○ No

3.1a Approximately how many (confirmed cases) ______________________
(By the date of completing this survey)

3.1b Which services were they in

Tick all that apply

- ☐ In-patient hospice / palliative care unit
- ☐ Home palliative care service
- ☐ Acute hospital
- ☐ Care home
- ☐ Other (a box will open below)

(Tick all that apply)

3.1c Please specify the other services ______________________

3.2 Have you had any patients with suspected (untested but with clinical diagnosis/symptoms) of COVID-19

(additional questions will open if you tick yes)

○ Yes ○ No

3.2a Approximately how many (suspected cases) (by the date of completing this survey) ______________________
(Must be a number)

3.2b Which services were they in

- ☐ In-patient hospice / palliative care unit
- ☐ Home palliative care service
- ☐ Acute hospital
- ☐ Care home
- ☐ Other (a box will open below)

(Tick all that apply)

3.2c Please specify the other services ______________________

3.2d Of the patients you have seen with suspected or confirmed COVID-19 would you say that they were

☐ Patients who are severely ill or dying due mainly to COVID-19
☐ Patients with pre-existing illnesses / co-morbidities as well as COVID-19 who are severely ill or dying
☐ Patients known to your service already who now have COVID-19
☐
(Tick all that apply)

3.3 Have you had any family members / close friends of your patients who had suspected or confirmed COVID-19

◯ Yes ◯ No

3.4 Have you had staff with suspected or confirmed COVID-19

(if yes - additional questions will open)

◯ Yes ◯ No

3.4a Were the staff

☐ Nurses
☐ Physicians
☐ Allied health professionals, managed
☐ Reception / administrative staff
☐ Managers
☐ Others (a box will open below)
(Tick all that apply)

3.4b Please specify the other staff

__________________________________

3.4c What impact has this had on your service

__________________________________

3.5 Have you had volunteers with suspected or confirmed COVID-19

(if yes - an additional question will open)

◯ Yes ◯ No

3.5a What impact has this had on your service

__________________________________

#### 4. HOW HAVE YOUR SERVICES CHANGED IN RESPONSE TO COVID-19

4.1 Have your services changed

◯ Yes ◯ No

4.2 Would you say overall you are more busy or less busy that before the COVID-19 Pandemic

◯ A lot more busy
◯ Slightly more busy
◯ About the same
◯ Slightly less busy
◯ Much less busy
(Tick the answer that best applies)

4.3 Why is this

__________________________________________

4.4 Have you lost staff from your service who have been moved to help the NHS elsewhere

(if yes - a box for details will open)

◯ Yes ◯ No

4.4a Please give details (lost staff)

__________________________________

4.5 Have you had staff offered to help your service from health services elsewhere | ◯ Yes ◯ No

(if yes - a box for details will open)

4.5a Please give details (offered staff) ________________________________

4.6 Have you changed how your staff work | ◯ Yes ◯ No

(if yes - a box for details will open)

4.6a Please give details (how work) ________________________________

4.7 Have you changed where your staff work (e.g. home working) | ◯ Yes ◯ No

(if yes - a box for details will open)

4.7a Please give details (where work) ________________________________

4.8 Have you changed how your volunteers engage and where | ◯ Yes ◯ No

(if yes - a box for details will open)

4.8a Please give details (changed volunteers) ________________________________

#### Use of virtual technologies

4.9 Would you say that you are using virtual technologies (e.g. zoom / teams etc.) with patients and families

◯ A lot more
◯ Slightly more
◯ About the same
◯ Slightly less
◯ Much less
(Tick the answer that best applies)

4.10 Would you say that you are using virtual technologies (e.g. zoom / teams etc.) with colleagues

◯ A lot more
◯ Slightly more
◯ About the same
◯ Slightly less
◯ Much less
(Tick the answer that best applies)

4.11 What have been the difficulties of using virtual technologies ________________________________

4.12 What has worked well when using virtual technologies ________________________________

#### Advance care planning

4.13 Would you say you are now involved directly with patients / families in advance care planning

◯ A lot more
◯ Slightly more
◯ About the same
◯ Slightly less
◯ Much less
(Tick the answer that best applies)

4.14 Would you say you are now involved advising / supporting others and / or educating about advance care planning

◯ A lot more
◯ Slightly more
◯ About the same
◯ Slightly less
◯ Much less
(Tick the answer that best applies)

4.15 In what ways (if any) have you changed how you are supporting advance care planning

__________________________________

4.16 What would you say are the main challenges for advance care planning during the COVID-19 pandemic

__________________________________

#### Bereavement support

4.17 Would you say that you provide more or less bereavement support than before

◯ A lot more
◯ Slightly more
◯ About the same
◯ Slightly less
◯ Much less
(Tick the answer that best applies)

4.18 What would you say are the main challenges for bereavement support during the COVID-19 pandemic

__________________________________

#### Other support

4.19 How are you supporting patients with COVID-19 who are from more disadvantaged sociodemographic communities (e.g. areas with greater poverty, poor housing, homelessness)

__________________________________

4.20 Have you encountered patients or families with COVID-19 who are from black and minority ethnic groups

◯ Yes ◯ No

(if yes - a box for details of differences will open)

4.20a Are there any differences in how you are supporting or reaching them

__________________________________

4.21 Are there any groups (e.g. with different religions, cultures) where you have found supporting the individual needs of people affected by COVID-19 is particularly challenging

__________________________________

#### Effects on patients who do not have COVID-19

4.22 How has COVID-19 changed how you are supporting the types of patients (e.g. with symptoms and progressive illness) that you would usually support

__________________________________

4.23 How has COVID-19 changed how you are supporting the families / those important to patients that you would usually support

__________________________________

#### 5. CHANGES IN SPECIFIC SERVICES, E.G. IN-PATIENT, HOSPICE, VOLUNTEERS

The next questions ask about some specific changes that might have occurred, please answer only the sections that apply to your services

5.1 Have there been changes in these areas

(additional questions will open for each choice)

- ☐ In-patient beds in your own service
- ☐ How you provide support for patients in acute hospitals
- ☐ How you provide support for patients in their own homes
- ☐ How you provide support for patients in care homes (including nursing homes)

(Tick all that apply)

#### Changes in in-patient beds in your service

5.1a What changes were there in how you used your beds (if any)

__________________________________

5.1b Number of beds

- ◯ Increased
- ◯ Stayed about the same
- ◯ Decreased

5.1c Any changes to admission criteria (if so what was the change)

__________________________________

5.1d Any changes to out of hours admissions (e.g. evenings / weekends - if so what was the change)

__________________________________

#### Changes in how you provide support for patients in acute hospitals

5.1e Numbers of patients needing support

- ◯ Increased
- ◯ Stayed about the same
- ◯ Decreased

5.1f Would you say your face to face contact with patients / their family members is in general

- ◯ A lot more
- ◯ Slightly more
- ◯ About the same
- ◯ Slightly less
- ◯ Much less

5.1g Would you say your face to face contact with staff is

◯ A lot more
◯ Slightly more
◯ About the same
◯ Slightly less
◯ Much less

5.1h Would you say your telephone / remote connection advice / support is

◯ A lot more
◯ Slightly more
◯ About the same
◯ Slightly less
◯ Much less

5.1i Have you changed how your team is organized (e.g. supporting patients with and without COVID-19) ________________________

5.1j Have you changed your working hours (and in what way) ________________________

5.1k Have you changed your working practices (and in what way) ________________________

#### Changes in how you provide support for patients in their own homes

5.1l Numbers of patients needing support

◯ Increased
◯ Stayed about the same
◯ Decreased

5.1m Would you say your face to face contact with patients / their family members is

◯ A lot more
◯ Slightly more
◯ About the same
◯ Slightly less
◯ Much less

5.1n Would you say your face to face contact with staff is

◯ A lot more
◯ Slightly more
◯ About the same
◯ Slightly less
◯ Much less

5.1o Would you say your telephone / remote connection advice / support is

◯ A lot more
◯ Slightly more
◯ About the same
◯ Slightly less
◯ Much less

5.1p Have you changed how your team is organized (e.g. supporting patients with and without COVID-19) ________________________

5.1q Have you changed your working hours (and in what way) ________________________

5.1r Have you changed your working practices (and in what way) ________________________

5.1s Have you changed how medicines are given in the community (e.g. who sets up syringe drivers / families administering medicines)

◯ Yes ◯ No

(if yes - a box for details will open)

5.1t Please give details (changed how medicines are given in the community) ________________________

#### Changes in how you provide support for patients in care homes (including nursing homes)

5.1u Numbers of patients needing support

◯ Increased
◯ Stayed about the same
◯ Decreased

5.1v Would you say your face to face contact with patients / their family members is

◯ A lot more
◯ Slightly more
◯ About the same
◯ Slightly less
◯ Much less

5.1w Would you say your face to face contact with staff is

◯ A lot more
◯ Slightly more
◯ About the same
◯ Slightly less
◯ Much less

5.1x Would you say your telephone / remote connection advice / support is

◯ A lot more
◯ Slightly more
◯ About the same
◯ Slightly less
◯ Much less

5.1y Have you changed how your team is organized (e.g. supporting patients with and without COVID-19)

__________________________________

5.1z Have you changed your working hours (and in what way)

__________________________________

5.1α Have you changed your working practices (and in what way)

__________________________________

#### Changes to how you are supporting families / those important to patients

5.2 How would you say you are supporting families / those important to patients compared to before

◯ A lot more
◯ Slightly more
◯ About the same
◯ Slightly less
◯ Much less

5.3 Have you changed how you contact and work with families / those important to patients

◯ Yes ◯ No
(If yes, a box for details will open)

5.3a Please give details (changed how contact and work with families)

__________________________________________

#### Changes to how you are deploying volunteers

5.4 How would you say you are deploying volunteers compared to before

◯ A lot more
◯ Slightly more
◯ About the same
◯ Slightly less
◯ Much less

5.5 Have you changed how you deploy volunteers ◯ Yes ◯ No

(if yes - a box for details will open)

5.5a Please give details (changed how contact and work with families) __________________________________

#### 6. CHALLENGES AND INNOVATIONS IN RESPONSE TO COVID-19

We want to know more about the challenges you have faced, their impacts on your services and care, how you have responded to them and what you found to be your successful innovations

Please tick the challenges that you have faced in your service during COVID-19 Pandemic during the past 1 month

6.1 Have you had problems accessing personal protective equipment ◯ Yes ◯ No

(if yes - additional questions will open)

6.1a Please specify what you had a shortage of __________________________________

6.1b What did you do about this __________________________________

6.1c Has this been a problem in the last 7 days
◯ Yes
◯ Sometimes
◯ No

6.2 Have you had a shortage of key medicines ◯ Yes ◯ No

(if yes - additional questions will open)

6.2a Please specify what you had a shortage of __________________________________

6.2b What did you do about this __________________________________

6.2c Has this been a problem in the last 7 days
◯ Yes
◯ Sometimes
◯ No

6.3 Have you had a shortage of other equipment (e.g. syringe drivers) ◯ Yes ◯ No

(if yes - additional questions will open)

6.3a Please specify what you had a shortage of __________________________________

6.3b What did you do about this __________________________________

6.3c Has this been a problem in the last 7 days

◯ Yes
◯ Sometimes
◯ No

6.4 Have you had a shortage of staff

◯ Yes ◯ No

(if yes - additional questions will open)

6.4a Please specify what you had a shortage of

__________________________________

6.4b What did you do about this

__________________________________

6.4c Has this been a problem in the last 7 days

◯ Yes
◯ Sometimes
◯ No

6.5 Have there been other effects on yourself and/or on staff that you think we should know about

__________________________________

6.6 Please tell us about any other challenges and whether or how you overcame them

__________________________________

6.7 What do you foresee will be the biggest challenges for COVID-19 in your service over the next 1-2 months

__________________________________

6.8 What would help you most to overcome these

__________________________________

---

**Now please tell us about your innovations. We are keen to learn what has worked best for you**

6.9 Please tell us about the change in practice or innovation that you think has been most successful to your working

__________________________________

6.10 Why is this

__________________________________

6.11 What would you say were the most important things that made this possible

__________________________________

6.12 Please list any other important changes / innovations you have made

__________________________________

#### 7. SYMPTOM MANAGEMENT

We are interested to learn more about how you are managing symptoms and psychological / emotional problems in our patients and the trajectories of care

##### How long are patients with COVID-19 under your palliative care service

7.1 What was the shortest time in hours ______________________________
(Give number of hours)

7.2 What was the longest time in days ______________________________
(Give number of days)

##### How are you managing symptoms

7.3 Please indicate which of these settings your management refers to (choose only one about which you have most experience)

- ◯ Inpatient hospital ward
- ◯ Inpatient hospital intensive care
- ◯ Community hospital
- ◯ Inpatient hospice / palliative care ward
- ◯ Community

##### The next questions ask about the treatments you are using

7.4 Do you have protocols for symptom management for COVID-19 patients

- ◯ Yes
- ◯ No
- ◯ Unsure

(Yes will ask for details)

7.4a What sources of information did you use

- ☐ Locally developed guidance
- ☐ NICE
- ☐ NHS
- ☐ Other (a box will open below)

7.4b Please specify the other protocols ______________________________

You can also email us your usual recommendations of any guidance you provide by email to: , marking the email CovPall in the subject line. We are still interested to know how well you find these treatments are working

##### Breathlessness

7.5 Which medicines and therapies do you usually prescribe

______________________________________

7.6 How effective do you find these e.g. time to give relief and how well it works

______________________________________

#### Agitation

7.7 Which medicines and therapies do you usually prescribe

__________________________________

7.8 How effective do you find these e.g. time to give relief and how well it works

__________________________________

#### Fever / Shivering

7.9 Which medicines and therapies do you usually prescribe

__________________________________

7.10 How effective do you find these e.g. time to give relief and how well it works

__________________________________

#### Cough

7.11 Which medicines and therapies do you usually prescribe

__________________________________

7.12 How effective do you find these e.g. time to give relief and how well it works

__________________________________

#### Pain

7.13 Which medicines and therapies do you usually prescribe

__________________________________

7.14 How effective do you find these e.g. time to give relief and how well it works

__________________________________

#### Other symptoms

7.15 Please provide details of the treatments you are using for any other symptoms you are seeing commonly in COVID-19, especially if this differs from usual palliative care practice

__________________________________

#### Additional comments

7.16 Please provide any additional comments you would like us to be aware of

__________________________________________

7.17 Please tell us if you would like us to help you by providing anything else, or any key questions that you think are important to answer

__________________________________________

#### Finally

7.18 Please indicate if you would like / are willing to be contacted regarding any of the following
(Tick all that apply)

☐ To receive copy of the early reports and our newsletters as the findings emerge
☐ For us to check any information with you
☐ To be acknowledged as responding to this questionnaire (listed along with other services) in the reports and any publications
☐ To participate in a subsequent survey similar to this one in 6-8 weeks time when you may have made more changes or had more experiences
☐ To collect pseudoanonymized data about a small series (around 10) of patients with COVID-19 in your service. This would involve collecting information on symptom severity, on first assessment in palliative care, in around 2 subsequent time points and at discharge or death, to understand more about the symptoms patients experience and their the effective treatments. This would not be a clinical trial, simply recording your practice and views

7.19 If you wish us to use a different Name or Email for the above (instead of the ones already given) please specify here

__________________________________________

7.20 How you would like your service acknowledged in any reports, if applicable

___________________________________

You will be free to opt out of receiving the updates at any time, your details will not be passed onto other organisations or used for anything other than with your explicit consent above. Your individual responses will remain confidential, they will be analysed pseudonymously by the research team, with your service identified only by a code number unless you explicitly ask us to do otherwise.

#### Thank you for your help at this difficult time

#### Appendix III: Acknowledgement of services

We would like to thank all the palliative care services and leads across the world for responding to the CovPall online survey.

The following persons and services indicated they were happy to be acknowledged for responding to the CovPall online survey:

| |
|---|
| Alexander Devine Children's Hospice Service |
| Alison Wardrop District Nursing Sister, Hywel Dda University Health Board |
| Ashgate Hospicecare |
| Bangalore Baptist Hospital Palliative Care Service |
| BCUHB West HSPCT |
| Beatson West of Scotland Cancer Centre HSPCT |
| Beaumond House Community Hospice |
| Betsi Cadwaladr University Health Board SPCT |
| Birmingham St Mary's Hospice |
| Bluebell Wood Children's Hospice |
| Bolton NHS Foundation Trust Specialist Palliative Care Team |
| Bury Community Specialist Palliative Care Team |
| Cambridge University Hospital NHS Foundation Trust Palliative Care Service |
| Carmarthenshire Specialist Palliative Care Service, Hywel Dda University Health Board |
| Center for Palliative Care, University Hospital Cologne, Germany |
| Cesta domů, home hospice |
| Chelsea and Westminster Hospital |
| CHFT Hospital SPCT |
| Children's hospices across Scotland (CHAS) |
| City Hospice Cardiff |
| Claire House Children's Hospice |
| Clinique de Médecine Palliative, CHU de Lille, France |
| CNWL UCLH |
| Community based clinical nurse specialists |
| Community Specialist Palliative Care Team, St Ann's Hospice, Salford |
| Consultant Dr Paul Coulter, CNS Elizabeth Anderson and CNS Kellyann O'Neill, Palliative Care Team, Inverclyde Royal Hospital |
| Cornwall Hospice Care |
| Croydon Health Services Macmillan Specialist Palliative Care Team |
| Cwm Taf Morgannwg SPCT North |
| Department of Palliative Care, Homerton University Hospital NHS Foundation Trust |
| Department of Palliative care, Sheffield Teaching Hospitals NHS Foundation Trust |
| Department of Palliative Medicine, LMU Munich, Germany |
| Department of Palliative Medicine, Southern Sector, South East Sydney Local Health District, New South Wales, Australia |
| Deutsche PalliativStiftung |
| Dorothy House Hospice |
| Dove House Hospice, Hull |
| Dr Charles Daniels, Medical Director, St Luke's Hospice Harrow and Consultant in Palliative Medicine, LNWHUT. |

| |
|---|
| Dr Jonathan Downie, Consultant in Paediatric Palliative Medicine, Supportive and Palliative Care Team, Royal Hospital for Children, Glasgow |
| East Anglia's Children's Hospices (EACH) |
| East Cheshire Hospice |
| East Sussex Healthcare NHS Trust Supportive and Palliative Care Team |
| Eastern Health |
| Ed Dubland MD CCFP pc |
| Ellenor Adult Services |
| Ellenor Children's Services |
| Ellenor Hospice, Coldharbour Road, Gravesend, Kent |
| Ellenor Inpatient Ward |
| Ellenor Wellbeing Service |
| Elvis J Miti for Community Palliative Care, UZIMA Project Ndanda, Mtwara, Tanzania. |
| ESNEFT specialist palliative care team |
| Family Support Team, Children's Hospices Across Scotland |
| Farleigh Hospice |
| Felicia Kontopidis, Journey Home Hospice |
| Fondazione Antea, Rome, Italy |
| Fondazione FARO, Turin, ITALY |
| Forest Holme Hospice, Poole Hospital NHS Foundation Trust |
| Forget Me Not Children's Hospice |
| Garden House Hospice Care |
| GHNHSFT Specialist Palliative Care Community team |
| Glasgow Royal Infirmary Hospital Specialist Palliative Care Team |
| Greenwich & Bexley Community Hospice |
| GSTT Community Palliative Care Team |
| Hambleton & Richmondshire SPCT |
| Hampshire Hospitals NHS Foundation Trust |
| Harrogate and District NHS Foundation Trust Palliative Care Team |
| Heart of Kent Hospice |
| Helen & Douglas House |
| Hospice De Wingerd (Charim Care Group) |
| Hospice Isle of Man |
| Hospice of St. Francis Berkhamsted, Herts |
| Hospice of The Good Shepherd, Chester |
| Hospice Waikato |
| Hospiscare Devon |
| Hospital Divina Providencia, El Salvador |
| Hospital Palliative Care Team, Kingston Hospital NHS Foundation Trust |
| Hospital Palliative Care Team, University Hospitals of North Midlands |
| Hospital Palliative Care team, University Hospital Southampton |
| Hospital Selayang Palliative Care Unit |

| |
|---|
| Hospital Specialist Palliative care team |
| Hospital Specialist Palliative Care Team, University Hospitals of Leicester NHS Trust |
| Hull University Teaching Hospitals NHS Trust |
| Instituto de Investigaciones Médicas Alfredo Lanari, Universidad de Buenos Aires |
| Isabel Hospice |
| J Yeomans, St Richards Hospice (Worcester) |
| James Cook University Hospital Acute SPCT |
| Thitima Phosri |
| John Taylor Hospice |
| Helen Hubert, Palliative care and End of Life Care Pharmacist, St Richard's Palliative Care Team |
| Katharine House Hospice Banbury |
| Kauniala Hospital |
| Kenelm F McCormick M.D., Akron, Ohio |
| Kilbryde Hospice, South Lanarkshire |
| Kim Jones, Deputy Head of Clinical Services at Hospice of the Valleys |
| King's College Hospital NHS Foundation Trust |
| Klinik Susenberg, Zürich |
| Knysna Sedgefield Hospice |
| Lanarkshire SPC service |
| Laois Offaly Palliative Care Service Cloneygowan Co Offaly |
| Leeds Teaching Hospitals NHS Trust Palliative Care Team |
| Limewood Dementia Service, Stafford |
| London Northwest Healthcare University Trust |
| LOROS Hospice, Leicester |
| Lynn Bushor, Department of Veteran's Affairs |
| Macmillan Unit / Royal Bournemouth & Christchurch |
| Manchester Foundation Trust - MRI |
| Manchester Foundation Trust WTWA |
| Maricruz Macias Montero, Hospital General de Segovia, Seccion de Geriatria. |
| Marie Curie Community Nursing Service London |
| Marie Curie Hospice, Cardiff and the Vale |
| Marie Curie Hospice West Midlands |
| Marie Curie Hospice, Hampstead |
| Marie Curie Services in Lothian |
| Martlets Hospice |
| Martlets Hospice, Brighton |
| Maura Farrell Miller, Director, Hospice and Palliative Care Program, Department of Veterans Affairs Medical Center, Florida |
| Mellannorrlands Hospice Sundsvall Sweden |
| Michael Sobell Hospice and Harlington Hospice |
| Mid Cheshire Hospital NHS Trust Specialist Palliative Care Team |
| Midhurst Macmillan Service (Sussex Community NHS Foundation Trust) |

| |
|---|
| Municipal Hospital Dr Cornel Igna, Palliative Care Department from Campia Turzii |
| Myton Hospice |
| Newham University Hospital Specialist Palliative Care Team - Barts Health NHS Trust |
| NHS Ayrshire & Arran Supportive Care Team |
| NHS Fife Specialist Palliative Care Service |
| NHS Grampian Specialist Palliative Care Team |
| NHS Specialist Palliative Care Unit |
| Nightingale House Hospice, North Wales |
| Ninewells Hospital Palliative Care Team |
| North London Hospice |
| Northern Ireland Hospice Adult Service |
| Northumbria Healthcare NHS Foundation Trust |
| Nottingham University Hospitals NHS Trust |
| Ofra Fried, Palliative Care Specialist, Townsville University Hospital, Queensland |
| Overgate Hospice |
| Palliativa Care Unit. west health Area. Valladolid. Castilla y León .Spain |
| Palliative and End of Life care Team, Newcastle upon Tyne Hospitals NHS Foundation Trust |
| Palliative Care Department, Bangalore Baptist Hospital |
| Palliative Care Service, Chaim Sheba Medical Center, Ramat-Gan, Israel |
| Palliative Care Service, Gloucestershire Hospitals NHS Foundation Trust |
| Palliative Care Team-Bassett Medical Center, Cooperstown NY |
| Palliative Care Team, Centre for Pain Management and Palliative Care, Haukeland University Hospital, Bergen, Norway |
| Palliative Care Unit, Bolzano, Italy |
| Palliative Care Unit, Bassini Hospital, Cinisello Balsamo, Milan, Italy |
| Palliative Care, Kantonsspital Olten, Switzerland |
| Palliative Medicine, UHDB NHS Trust |
| Palliaviva |
| Peace Hospice Care |
| Pembrokeshire Specialist Palliative Care service, Hywel Dda University Health Board |
| Phyllis Tuckwell Hospice Care |
| Pilgrims Hospices in East Kent |
| Pippa Hawley, Medical Director, Pain and Symptom Management Palliative Care, BC Cancer. |
| Portsmouth Hospital NHS Trust Palliative Care Service |
| Princess Alice Hospice |
| Prof Olaitan A Soyannwo |
| Queen Elizabeth University Hospital Glasgow Specialist Palliative Care Team, and Prince and Princess of Wales Hospice Glasgow |
| Queenscourt Supportive & Specialist Palliative Care Service |
| Rotherham NHS Foundation Trust Palliative Care Team |
| Rowans Hospice |
| Rowcroft Hospice |

| |
|---|
| Royal Berkshire Foundation Trust Hospital Palliative Care Team |
| Royal Trinity Hospice |
| Sabar Health Home Hospital |
| Saint Francis Hospice |
| Salford Royal Hospital Specialist Palliative Care Team |
| Salisbury Specialist Palliative Care service |
| SAPV des OSP Karlsruhe |
| SCCU SEVILLA SUR, SAS. SPAIN. |
| Sengkang General Hospital, Singapore |
| Shooting Star Children's Hospice |
| Sobell House, Oxford University Hospitals |
| South East Palliative Care Services |
| South East Palliative Care, University Hospital Waterford, Ireland |
| South Tees Community Specialist Palliative Care MRC |
| South Tipperary Hospice Homecare Team |
| South Tyneside District Hospital Specialist Palliative Care Team, South Tyneside and Sunderland NHS Foundation Trust |
| Specialist Palliative Care Hospital Team. East Kent University Foundation Trust |
| Specialist Palliative Care Service South Eastern Health and Social Care Trust |
| Specialist Palliative Care Team Imperial College Healthcare NHS Trust |
| Specialist Palliative Care Team, Coventry and Warwickshire Partnership NHS Trust |
| Specialist Palliative Care Team, West Middlesex University Hospital, Chelsea and Westminster NHSFT |
| Specialist Palliative Care Service, County Durham and Darlington NHS Foundation Trust |
| St Ann's Hospice |
| St Barnabas Hospice, Lincolnshire |
| St Barnabas House, Worthing |
| St Bartholomew's Hospital, Barts Health |
| St Catherine's Hospice, Preston |
| St Christopher's |
| St Clare Hospice, Essex |
| St Columba's Hospice, Edinburgh |
| St Gemma's Hospice - Leeds, West Yorkshire. |
| St Helena |
| St John of God Murdoch Hospital |
| St John's Hospice, Doncaster, RDASH |
| St Johns Hospice, Lancaster |
| St Leonard's Hospice, York |
| St Luke's Cheshire Hospice |
| St Luke's Combined Hospices, Cape Town, South Africa |
| St Luke's Hospice, Sheffield. |
| St Luke's Hospice, Kenton, Harrow covering London boroughs of Brent and Harrow. Ursula Reeve, Director of Patient Services. Cathy Hanrott, Hospice Service Navigator |
| St Margaret's Hospice, Somerset. |

| |
|---|
| St Martha's Hospital Palliative Care Team |
| St Mary's Hospice, Ulverston, Cumbria |
| St Michael's Hospice, Hastings and Rother |
| St Oswald's Hospice, Newcastle |
| St Peter's Hospice |
| St Richards Hospice |
| St Wilfrid's Hospice, Chichester |
| St Wilfrid's Hospice, Eastbourne. |
| St. Luke's Hospice Community Services South West Essex |
| Stockport Specialist Palliative Care Service |
| Strathcarron Hospice |
| Sue Ryder Leckhampton Court Hospice |
| Sue Ryder Manorlands Hospice |
| Susanna Sandöy, Capio Asih Nacka, |
| Swansea Bay University Health Board Specialist Palliative Care Service (which includes Ty Olwen Hospice) |
| Symptom Control and Palliative Care Team, Royal Marsden NHS Foundation Trust |
| Tatiana Chavouzi |
| Tayside Palliative Care Service |
| The Arthur Rank Hospice, Cambridge |
| The Camden, Islington ELiPSe Palliative Care Team |
| The Hillingdon Hospital Palliative Care team |
| The Prince of Wales Hospice, Pontefract |
| The Wisdom Hospice and Medway Community Healthcare |
| TOPAT Zuyderland MC |
| Trinity Hospice & Brian House Children's Hospice |
| Trinity Hospice, Blackpool and Blackpool Teaching Hospitals |
| Tynedale Hospice at Home |
| Unità di Cure Palliative - Hospice Casale Monferrato ASL AL Piemonte |
| University Hospital Lewisham Macmillan Specialist Palliative Care Team, Lewisham and Greenwich NHS Trust |
| University Teaching Hospital Lusaka Adult Medicine Palliative Care Unit |
| Waterford Hospice Homecare Team |
| Weldmar Hospicecare |
| West Suffolk Hospital NHS Foundation Trust |
| Western Australia Paediatric & Adolescent Palliative Oncology, Perth Children's Hospital |
| Weston Hospicecare |
| Whittington Health Palliative Care Service |
| Wigan & Leigh Hospice |
| Wrighinton, Wigan and Leigh NHS Teaching Trust (Supportive and Palliative Care Team) |
| York Teaching Hospital Foundation Trust |
| York Teaching Hospitals |
| Zuercher Lighthouse |

| |
|---|
| Zuzana Kremenova |
