## Supplementary material for "The challenges of caring for people dying from COVID-19: a multinational, observational study of palliative and hospice services (CovPall)": CHERRIES, Checklist of MORECare Statement and STROBE Checklist

### CovPall survey, checklist for Reporting Results of Internet E-Surveys (CHERRIES)

| | **Checklist for Reporting Results of Internet E-Surveys (CHERRIES)** | | |
|---|---|---|---|
| ***Item Category*** | ***Checklist Item*** | ***Description*** | ***PAGE*** |
| **Design** | | | |
| | Describe survey design | Services were identified and contacted through national and multinational palliative care and hospice organisations and provided with a link to complete an on-line survey. | 5 |
| **IRB (Institutional Review Board) approval and informed consent process** | | | |
| | IRB approval | The survey received ethical (Institutional Review Board) approval from King's College London Research Ethics committee (LRS-19/20-18541) | 4 |
| | Informed consent | Services were identified and contacted through national and multinational palliative care and hospice organisations and provided with a link to an on-line survey to be completed by the medical or nursing lead or their nominee. Completion indicated consent. The email attached the participant information sheet with details of the study rationale, ethical approval, data protection and management, investigators and contacts for further information or concern. It was intended to be brief, taking around 30 minutes to complete. Free-text explanatory comments were invited. | 5 |
| | Data protection | We developed and piloted a secure, password-protected web-based data entry portal (in the Research Electronic Data Capture (REDCap) at the study coordinating centre, King's College London. Services could keep their identity hidden if they wished, but most chose to provide an email for contact. Data were anonymised before analysis. | 5, Supplementary file Box S1 |
| **Development and pre-testing** | | | |
| | Development and testing | The questionnaire was developed and piloted by the CovPall study team building on an earlier survey of Italian hospices, adding questions on the impact of and response to COVID-19. The study team developed and piloted a secure, password-protected web-based data entry portal (in the Research Electronic Data Capture (REDCap) at the study coordinating centre, King's College London. | 5, Supplementary file Box S1 |
| **Recruitment process and description of the sample having access to the questionnaire** | | | |
| | Open survey versus closed survey | This was a closed survey. Services were identified and contacted through national and multinational gatekeeper palliative care and hospice organisations and asked that their medical or nursing lead, or their nominee, complete an on-line survey available via a link. The study team developed and piloted a secure, password-protected web-based data entry portal (in the Research Electronic Data Capture (REDCap) at the study coordinating centre, King's College London. | 5, Supplementary file Box S1 |
| | Contact mode | Services were identified and contacted through national and multinational gatekeeper palliative care and hospice organisations | 5, Supplementary file Box S1 |
| | Advertising the survey | Services were identified and contacted through national and multinational gatekeeper palliative care and hospice organisations. The email attached the participant information sheet with details of the study rationale, ethical approval, data protection and management, investigators and contacts for further information or concern. No incentives were offered for completion. The CovPall study was presented at relevant on-line meetings and discussed with gatekeeper organisations | 5, Supplementary file Box S1 |

| | **Checklist for Reporting Results of Internet E-Surveys (CHERRIES)** | | |
|---|---|---|---|
| ***Item Category*** | ***Checklist Item*** | ***Description*** | ***PAGE*** |
| | | and in blogs, to inform the methods, questions and to raise awareness and engagement. | |
| **Survey administration** | | | |
| | Web/E-mail | Services were identified and contacted through national and multinational gatekeeper palliative care and hospice organisations and asked that their medical or nursing lead, or their nominee, complete an on-line survey available via a link. The study team developed and piloted a secure, password-protected web-based data entry portal (in the Research Electronic Data Capture (REDCap) at the study coordinating centre, King's College London. | 5, Suppleme ntary file Box S1 |
| | Context | As stated above, services were identified and contacted through national and multinational gatekeeper palliative care and hospice organisations and asked that their medical or nursing lead, or their nominee, complete an on-line survey available via a link. | 5, Suppleme ntary file Box S1 |
| | Mandatory/voluntary | It was a voluntary survey as service leads were not mandated to complete it. | Suppleme ntary file, appendix II |
| | Incentives | There was no incentive offered for survey completion | Suppleme ntary file Box S1 |
| | Time/Date | The survey opened on April 23rd and closed July 31st 2020 | Suppleme ntary file Box S1 |
| | Randomization of items or questionnaires | Items were not randomised or alternated. | Suppleme ntary file, appendix II |
| | Adaptive questioning | We used adaptive questioning in order to reduce the number and complexity of questions. | Suppleme ntary file, appendix II |
| | Number of Items | We had an average of 12 questionnaire items per page | Suppleme ntary file, appendix II |
| | Number of screens (pages) | 16 pages | Suppleme ntary file, appendix II |
| | Completeness check | The research team audited the data weekly to ensure data entry completeness and sent monthly missing data and incomplete entry reports to the research associates and administrators, and where consent permitted to relevant respondents to check validity. | Suppleme ntary file Box S1 |
| | Review step | Respondents could review and change their answers before submitting the survey. They could also pause the questionnaire by clicking the "Save and Return Later" button at the bottom of each page. There were given a code to enable them continue later. There was a "returning" option at the top right of the page. If respondents wanted to correct any errors after submitting the survey, they were informed that they could email the research team at with CovPall in the subject line | Suppleme ntary file, appendix II |
| **Response rates** | | | |

| | **Checklist for Reporting Results of Internet E-Surveys (CHERRIES)** | | |
|---|---|---|---|
| ***Item Category*** | ***Checklist Item*** | ***Description*** | ***PAGE*** |
| | Unique site visitor | In total 489 questionnaire were commenced, 477 completed (completion rate 97·5%); of these 15 were duplicates and 2 triplicates of entries with the same name/email; 2 were invalid being from one researcher without a palliative care service, leaving 458 valid responses. We were able to determine this through the email addresses provided. | 6 |
| | View rate (Ratio of unique survey visitors/unique site visitors) | Not applicable. Respondents were invited to participate through national and multinational gatekeeper palliative care and hospice organisations | 5 |
| | Participation rate (Ratio of unique visitors who agreed to participate/unique first survey page visitors) | Not applicable. Respondents were invited to participate through national and multinational gatekeeper palliative care and hospice organisations | 5 |
| | Completion rate (Ratio of users who finished the survey/users who agreed to participate) | In total 489 questionnaire were commenced, 477 completed (completion rate 97·5%); of these 15 were duplicates and 2 triplicates of entries with the same name/email; 2 were invalid being from one researcher without a palliative care service, leaving 458 valid responses. | 6 |
| **Preventing multiple entries from the same individual** | | | |
| | Cookies used | Cookies were not used. In total 489 questionnaire were commenced, 477 completed (completion rate 97·5%); of these 15 were duplicates and 2 triplicates of entries with the same name/email; 2 were invalid being from one researcher without a palliative care service, leaving 458 valid responses | 6 |
| | IP check | IP check was not carried out. However, as stated above, the unique email addresses were used to identify duplicate entries. Respondents with duplicate entries were contacted and asked which entry should be included in the analysis. | 6 |
| | Log file analysis | As stated above, the unique email addresses were used to identify duplicate entries. Respondents with duplicate entries were contacted and asked which entry should be included in the analysis. | 6 |
| | Registration | As stated above, the unique email addresses were used to identify duplicate entries. Respondents with duplicate entries were contacted and asked which entry should be included in the analysis. | 6 |
| **Analysis** | | | |
| | Handling of incomplete questionnaires | After removing duplicates and ineligible entries all available data were analysed. Missing data were not imputed. | 5 |
| | Questionnaires submitted with an atypical timestamp | Not applicable. After removing duplicates and ineligible entries all available data were analysed. | 5 |
| | Statistical correction | Not applicable | |

### STROBE Statement—Checklist of items that should be included in reports of *cross-sectional studies*

| | Item No | Recommendation | |
|---|---|---|---|
| **Title and abstract** | 1 | (*a*) Indicate the study's design with a commonly used term in the title or the abstract | Yes |
| | | (*b*) Provide in the abstract an informative and balanced summary of what was done and what was found | Yes |
| **Introduction** | | | |
| Background/rationale | 2 | Explain the scientific background and rationale for the investigation being reported | Yes |
| Objectives | 3 | State specific objectives, including any prespecified hypotheses | Yes |
| **Methods** | | | |
| Study design | 4 | Present key elements of study design early in the paper | Yes |
| Setting | 5 | Describe the setting, locations, and relevant dates, including periods of recruitment, exposure, follow-up, and data collection | Survey opened on the 23rd of April 2020 and closed on the 31st of July 2020 |
| Participants | 6 | (*a*) Give the eligibility criteria, and the sources and methods of selection of participants | Yes |
| Variables | 7 | Clearly define all outcomes, exposures, predictors, potential confounders, and effect modifiers. Give diagnostic criteria, if applicable | Yes |
| Data sources/ measurement | 8* | For each variable of interest, give sources of data and details of methods of assessment (measurement). Describe comparability of assessment methods if there is more than one group | Yes, textual and quantitative variables described |
| Bias | 9 | Describe any efforts to address potential sources of bias | Yes – Page 5 Data were anonymised before analysis |
| Study size | 10 | Explain how the study size was arrived at | Yes – page 5 |
| Quantitative variables | 11 | Explain how quantitative variables were handled in the analyses. If applicable, describe which groupings were chosen and why | We used contingency tables, $\chi 2$ tests, and correlations in SPSS (v26) and then multivariable logistic regression to explore relationships between variables was done in STATA (v16) |
| Statistical methods | 12 | (*a*) Describe all statistical methods, including those used to control for confounding | We used contingency tables, $\chi 2$ tests, and correlations in SPSS (v26) and then multivariable logistic regression to explore relationships between variables was done in STATA (v16) |
| | | (*b*) Describe any methods used to examine subgroups and interactions | Not applicable |
| | | (*c*) Explain how missing data were addressed | Missing data were not imputed due to |

| | | | |
|---|---|---|---|
| | | | limitations inherent in the commonly used approaches for handling missing data |
| | | (*d*) If applicable, describe analytical methods taking account of sampling strategy | Not applicable |
| | | (*e*) Describe any sensitivity analyses | Not applicable |
| **Results** | | | |
| Participants | 13* | (a) Report numbers of individuals at each stage of study—eg numbers potentially eligible, examined for eligibility, confirmed eligible, included in the study, completing follow-up, and analysed | The numbers potentially eligible could not be determined because this was an online survey. However, a total of 489 questionnaire were commenced, and 477 completed (completion rate 97·5%); of these 15 were duplicates and 2 triplicates of entries with the same name/email; 2 were invalid being from one researcher without a palliative care service, leaving 458 valid responses |
| | | (b) Give reasons for non-participation at each stage | N/A |
| | | (c) Consider use of a flow diagram | N/A |
| Descriptive data | 14* | (a) Give characteristics of study participants (eg demographic, clinical, social) and information on exposures and potential confounders | Yes – see table 1 and supplementary file I, table S1 for full details |
| | | (b) Indicate number of participants with missing data for each variable of interest | Yes – see table 1 and supplementary file I, table S1 for full details |
| Outcome data | 15* | Report numbers of outcome events or summary measures | Yes – see appendix table |
| Main results | 16 | (*a*) Give unadjusted estimates and, if applicable, confounder-adjusted estimates and their precision (eg, 95% confidence interval). Make clear which confounders were adjusted for and why they were included | See Tables 2 for the multivariate regression analysis. We preselected four dependent variables, presence or not of shortages of: personal |

| | |
|---|---|
| | protective equipment (PPE), staff, medicines, or other equipment (such as syringe drivers). Independent variables were: country/region, charitable or public management, settings (comprising four settings), experiences with COVID-19 and level of busyness. For each of the four multiple regression analyses we included independent variables showing $p<0·10$ in univariable analysis, excluding those exhibiting collinearity with independent variables already included if there was variance inflation factor >10 or chi-square test, $p < 0·05$. |
| (*b*) Report category boundaries when continuous variables were categorized | N/A |

| | | | |
|---|---|---|---|
| | | (*c*) If relevant, consider translating estimates of relative risk into absolute risk for a meaningful time period | N/A |
| Other analyses | 17 | Report other analyses done—eg analyses of subgroups and interactions, and sensitivity analyses | Yes<br>Page 5 (Free text responses) |
| **Discussion** | | | |
| Key results | 18 | Summarise key results with reference to study objectives | Yes<br>Pages 10-11 |
| Limitations | 19 | Discuss limitations of the study, taking into account sources of potential bias or imprecision. Discuss both direction and magnitude of any potential bias | Yes |
| Interpretation | 20 | Give a cautious overall interpretation of results considering objectives, limitations, multiplicity of analyses, results from similar studies, and other relevant evidence | Yes |
| Generalisability | 21 | Discuss the generalisability (external validity) of the study results | Yes |
| **Other information** | | | |
| Funding | 22 | Give the source of funding and the role of the funders for the present study and, if applicable, for the original study on which the present article is based | Yes – pages 2, 6 |

*Give information separately for exposed and unexposed groups.

**Note:** An Explanation and Elaboration article discusses each checklist item and gives methodological background and published examples of transparent reporting. The STROBE checklist is best used in conjunction with this article (freely available on the Web sites of PLoS Medicine at http://www.plosmedicine.org/, Annals of Internal Medicine at http://www.annals.org/, and Epidemiology at http://www.epidem.com/). Information on the STROBE Initiative is available at www.strobe-statement.org.

#### Checklist of MORECare Statement

| Category | Checklist items | Answer |
|---|---|---|
| Introduction/background | 1. Present theoretical framework for the intervention and levels of need established | Not applicable |
| | 2. Present objectives appropriate to the level of intervention development | Not applicable |
| Study design | 3. Indicate and justify stage in MRC guidance for development and evaluation of complex interventions, for example, feasibility, preliminary evaluation, efficacy/cost effectiveness and wider effectiveness | Not applicable |
| | 4. Feasibility stages should test both feasibility of the intervention and of methods of evaluation, including outcome measurement | Not applicable |
| | 5. Justify methods, considering appropriate use of existing data sets and secondary analysis as these may produce rapid information | Yes<br><br>Pages 4-5 |
| | 6. Justify methods of empirical studies considering mixed methods, observational studies and randomised trials | Yes<br><br>Pages 4-5 |
| Study team | 7. Ensure involvement from: (i) consumers, patients and caregivers; (ii) relevant clinicians; (iii) relevant methodologists to develop study questions, questionnaires and procedures; and (iv) researchers familiar with the challenges in EoLC studies | Yes. This research developed in response to a recent consultation with our existing patient and public involvement and engagement (PPIE) networks. Our study team and steering group includes clinicians and experts in the palliative care. |
| | 8. Ideally, involvement should be well established and continuing, beyond a specific study, with joint meetings or rotations between clinical and research staff | .We have weekly CovPall Team meetings that include clinical and research staff. We also have wider meetings and engagement with services through Hospice UK ECHO meetings, where we attend and regularly present findings. |
| Ethics | 9. Note in ethics committee application MORECare recommendations that it is ethically desirable for patients and families in EoLC to be offered involvement in research and MORECare evidence of patient willingness to be approached | Participants surveyed about their services were palliative care service leads. The research proposal developed in response to a recent consultation with our existing patient and public involvement and engagement (PPIE) networks. We received >40 responses via telephone, email and our online forum (www.csipublicinvolvement.co.uk). These identified the challenges for patients, their families, and members of the public, in relation to palliative and end of life care during the COVID-19 outbreak. |
| | | Not applicable |

| | | |
|---|---|---|
| | 10. Work within legal frameworks on mental capacity, consent and so on, to ensure that those who may benefit from interventions are offered an opportunity to participate if they wish | |
| | 11. Collaborate with patients and caregivers in the design of the study, vocabulary used in explaining the study, consent procedures and any ethical aspects | The research proposal developed in response to a recent consultation with our existing patient and public involvement and engagement (PPIE) networks. We received >40 responses via telephone, email and our online forum (www.csipublicinvolvement.co.uk). These identified the challenges for patients, their families, and members of the public, in relation to palliative and end of life care during the COVID-19 outbreak. Two patient and public involvement (PPI) members are part of our steering group. |
| | 12. Attend the ethics committee meeting with a caregiver or patient, as a means to help the committee better understand the patient perspective | We did not attend the ethics committee meeting with a caregiver or patient. However, patients and caregivers perspectives informed this research. |
| | 13. Ensure proportionality in patient and caregiver information sheets, appropriate to the study design and level of risk, as excessive information in itself can be tiring/distressing for very ill individuals | Not applicable |
| Participants | 14. Adjust eligibility criteria to recruit those patients who may benefit most from intervention, ensuring equipoise | Not applicable |
| Procedures | 15. Minimise burden for existing clinical staff for participation in the study | We minimised the burden of participation in this survey for palliative care leads. The palliative care providers were allowed to choose how they would like to provide data to minimise the burden to them. They had three options:<br>(a) to themselves enter data directly online into the bespoke data base, at a time of their choosing,<br>(b) if they preferred, they could give the information to a trained interviewer over the telephone or virtual connection (e.g. via Microsoft teams or Zoom) who entered the data for them, or<br>(c) if they preferred, be sent the survey as a word document via email to complete and return electronically (e.g. via secure NHS email).<br><br>There was flexibility around data entry with palliative care providers being able to enter data within 14-weeks. |
| | 16. Clearly distinguish between service received and research activity interviews in study arms when multiple interviews with patients are undertaken in trials, for example, using a graphical system | Not applicable |
| Outcome measures | 17. Choose outcome measures that meet the following criteria:<br>• established validity and reliability in relevant population<br>• responsive to change over time<br>• capture clinically important data<br>• easy to administer and interpret (for example, short and with low level of complexity)<br>• applicable across care settings to capture change in outcomes by location (for example, patients' home, hospital, hospice)<br>• able to be integrated into clinical care | Not applicable to survey |

| | | |
|---|---|---|
| | • minimise problems of response shift | |
| | 18. Consider including patients' experience of care, as this is central to many interventions | Not applicable |
| | 19. Select time points of outcome measurement to balance the value of early recording, to reduce attrition, but to allow enough time for the intervention to have had an effect | Not applicable |
| | 20. Consider the potential effect of response shift (that is, a change in a person's internal conceptualisation or calibration of the aspects measured). Questionnaires that include anchor points or descriptions of each response category may be less problematic in this regard | This has been considered and anchor time points and descriptions of response category are provided. |
| Missing data and attrition considerations | 21. Estimate in advance levels of, and reasons for, attrition and missing data, integrating these into sample size estimates and planned collection of data from proxies | Yes – page 5<br><br>After removing duplicate entries and ineligible entries (blank entries or where individuals indicated they were not from a palliative care service) all available data during the study period were analysed. Missing data were not imputed due to limitations inherent in the commonly used approaches for handling missing data. |
| | 22. Monitor during the study and report all levels of, and reasons for, attrition and other missing data | Yes – page 5<br>We could not report reasons for missing data in this survey. |
| | 23. Assume missing quantitative data NOT to be at random unless proven otherwise | Given the small number of missing data, we did not explore whether data were missing at random or not. Furthermore, missing data were not imputed due to limitations inherent in the commonly used approaches for handling missing data. |
| | 24. Test results from different methods of imputation – noting that 'using only complete cases' is a form of imputation | We used only complete cases in the analysis. |
| | 25. Use the MORECARE classification of attrition to describe causes of attrition: that is,<br><br>• ADD – attrition due to death;<br>• ADI - attrition due to illness;<br><br>AaR - attrition at random. | Not applicable |
| | 26. Consider reasons for missing data which are not due to attrition, for example missed questionnaire, or missed data item in questionnaire. Consider these in analysis and the potential imputations | While it is possible that some of survey respondents missed data items on the questionnaire, we did not explore this. We acknowledged the presence of missing data as a study limitation. |
| | 27. Mixed methods can be appropriate in all phases of development and evaluation | This study was not a mixed methods study. It was a multinational online survey of clinical leads of palliative care services. Free-text explanatory comments were also invited |

| | | |
|---|---|---|
| Mixed method studies | 28. Ensure appropriate multi-disciplinary skills mix or training of team | The CovPall team is multidisciplinary including quantitative and qualitative researchers, mixed methods researchers as well as people with different clinical backgrounds. The team includes doctors, nurses, a physiotherapist, and a pharmacist |
| | 29. Define the theoretical paradigm and method of integrating results and safeguards to ensure rigour at the outset | Not applicable |
| | 30. Plan investigation to avoid undue burden of qualitative and quantitative questionnaires – perhaps dividing data collection or selecting questions and/or sampling appropriately | Not applicable |
| | 31. Take into account any potential therapeutic effect of qualitative interviews where participants can express their feelings, if these are similar to components of the intervention | Not applicable |
| | 32. Ensure that those collecting data are appropriately trained in qualitative data collection | Not applicable |
| Implementation | 33. Consider implementation implications, including workforce and training needs, in all phases of the study | Not applicable |
| | 34. Integrate into preliminary evaluations and test feasibility of methods | Not applicable |
| Cost-effectiveness | 35. Collect data on use of services including health, voluntary, social and informal care, to take societal approach to care costs | Not applicable |
| | 36. Justify appropriate outcome measures to generate cost effectiveness | Not applicable |
